# SARS-CoV-2, influenza A/B and respiratory syncytial virus positivity and association with influenza-like illness and self-reported symptoms, over the 2022/23 winter season in the UK: a longitudinal surveillance cohort

**DOI:** 10.1101/2023.10.11.23296866

**Authors:** Elisabeth Dietz, Emma Pritchard, Koen Pouwels, Muhammad Ehsaan, Joshua Blake, Charlotte Gaughan, Eric Haduli, Hugh Boothe, Karina-Doris Vihta, Tim Peto, Nicole Stoesser, Philippa Matthews, Nick Taylor, Ian Diamond, Ruth Studley, Emma Rourke, Paul Birrell, Daniela De Angelis, Tom Fowler, Conall Watson, David Eyre, Thomas House, Ann Sarah Walker

## Abstract

**Background:** Syndromic surveillance often relies on patients presenting to healthcare. Community cohorts, although more challenging to recruit, could provide additional population-wide insights, particularly with SARS-CoV-2 co-circulating with other respiratory viruses.

**Methods:** We estimated positivity and incidence of SARS-CoV-2, influenza A/B, and RSV, and trends in self-reported symptoms including influenza-like illness (ILI), over the 2022/23 winter season in a broadly representative UK community cohort (COVID-19 Infection Survey), using negative-binomial generalised additive models. We estimated associations between test positivity and each of symptoms and influenza vaccination, using adjusted logistic and multinomial models.

**Findings:** Swabs taken at 32,937/1,352,979 (2.4%) assessments tested positive for SARS-CoV-2, 181/14,939 (1.2%) for RSV and 130/14,939 (0.9%) for influenza A/B, varying by age over time. Positivity and incidence peaks were earliest for RSV, then influenza A/B, then SARS-CoV-2, and were highest for RSV in the youngest and for SARS-CoV-2 in the oldest age-groups. Many test-positives did not report key symptoms: middle-aged participants were generally more symptomatic than older or younger participants, but still only ∼25% reported ILI-WHO and ∼60% ILI-ECDC. Most symptomatic participants did not test positive for any of the three viruses. Influenza A/B-positivity was lower in participants reporting influenza vaccination in the current and previous seasons (odds ratio=0.55 (95% CI 0.32,0.95)) versus neither season.

**Interpretation:** Symptom profiles varied little by aetiology, making distinguishing SARS-CoV-2, influenza and RSV using symptoms challenging. Most symptoms were not explained by these viruses, indicating the importance of other pathogens in syndromic surveillance. Influenza vaccination was associated with lower rates of community influenza test positivity.

**Funding:** UK Health Security Agency, Department of Health and Social Care, National Institute for Health Research.

## Introduction

Influenza and other respiratory illnesses place large burdens on patients and healthcare.^1,2^ Understanding within-season dynamics is critical to healthcare preparedness and vaccination planning. Routine syndromic and laboratory surveillance is commonly conducted using patients attending community doctors, hospitals, and ambulance services,^3^ thus being skewed towards symptomatic and more severe cases, and influenced by differential health-care seeking behaviours,^4^ and underestimating the community burden of seasonal influenza, as most cases are mild and/or asymptomatic.^5^ Alternative data sources include community surveys, e.g. the UK’s online participatory surveillance system ‘Flusurvey’.^6^ While such cohorts may provide better population-wide estimates, including mild illness, they may still not be representative, tending to underrepresent young children and older adults, both with higher risks of respiratory illness and distinct symptom patterns.^7,8^ Another challenge is the reliance on indicators such as influenza-like illness (ILI) in the absence of virological confirmation.^9^ The relationship between ILI and influenza positivity remains complex, influenced by differing case definitions,^10,11^ changes in co-circulation of other viruses (notably respiratory syntactical virus (RSV) and SARS-CoV-2) across seasons,^12,13^ age-specific dynamics,^14^ and the non-specific nature of influenza symptoms.^15,16^

Various studies have attempted to assess these relationships, but most have limited their scope to clinical settings, and/or focussed solely on influenza, and/or restricted to patients already reporting ILI or Acute Respiratory Illness (ARI).^7-9,12-14,17,18^ Similarly, influenza vaccine effectiveness evaluation typically uses disease endpoints, rather than protection from infection.^19^ Here we use the Office of National Statistics COVID-19 Infection Survey (CIS) to investigate the relationship between respiratory infection test positivity and ILI/other self-reported symptoms. This survey differs from sentinel laboratory surveillance in that routine nose and throat swab testing for SARS-CoV-2 (and on a smaller sub-sample, also for influenza A/B and RSV) was conducted on a community cohort, approached at random from address lists, not limited to those contacting healthcare services or with specific case presentations. We estimated SARS-CoV-2, influenza and RSV positivity and incidence across the 2022/2023 winter season, assessed associations between specific symptoms and test positivity and evaluated the effects of influenza vaccination on positivity.

## Methods

CIS was a large longitudinal household survey, broadly representative of the wider UK population (**Supplementary Methods**,^20^), conducting PCR tests for SARS-CoV-2 on self-collected nose and throat swabs and collecting questionnaire data including demographics and symptoms approximately monthly (**Supplementary Methods**,^21^). The study received ethical approval from the South Central Berkshire B Research Ethics Committee (20/SC/0195). From October 2022, a random subset of ∼750 swabs received per week were additionally tested by multiplex PCR (ThermoFisher TaqPath™ COVID-19, Flu A/B, RSV ComboKit) in a respiratory pilot study.^22^ We analysed swabs taken from 10-October-2022 to 26-February-2023 (≥350 respiratory pilot samples/week; ≤40 pilot samples/week outside this), when all survey assessments were conducted remotely, either online or by telephone, with swab kits posted to participants and returned by post/courier.

Each month, participants were asked whether they had experienced specific symptoms during the last seven days.^23^ This analysis included 12 symptoms solicited from the survey start (cough, sore throat, loss of taste, loss of smell, shortness of breath, fever, muscle ache (myalgia), weakness/tiredness (fatigue), headache, nausea/vomiting, abdominal pain, and diarrhoea) and four added September 2021 (wheezing, sneezing, ‘more trouble sleeping than usual’, and ‘loss of appetite or eating less than usual’), but excluded seven unrelated to respiratory illness added January 2022. Influenza-like illness (ILI) was defined using World Health Organisation (WHO) (concurrent fever and cough)^24^ and

European Centre for Disease Prevention and Control (ECDC) (co-presence of ≥1 respiratory symptom (cough, sore throat, shortness of breath) and ≥1 systemic symptom (fever, fatigue, headache, myalgia))^25^ definitions.

### Statistical methods

Percentage reporting different symptoms including ILI, and positivity for SARS-CoV-2 (full sample) and influenza A/B and RSV (respiratory pilot only), were estimated using negative binomial (log link) Generative Additive Models (GAM) (R *mgcv* package^26^), with a single explanatory variable for calendar time in days modelled with thin plate splines penalised on the third derivative^27^ with k=45 basis functions determining smoothness (approximately total study days(140)/3). Given expected variation, full sample models were run separately for six age-groups (2-6SY (school-year, ∼11y, **Supplementary Methods**), 7SY-11SY, 12SY-34, 35-49, 50-64, and 65+), collapsing to three wider age-groups (2-11SY, 12SY-49, 50+) for the smaller respiratory pilot.

Incidence was estimated using Richardson-Lucy-type deconvolution from daily estimates of test positivity and the distribution of infection (PCR positivity) duration^28,29^ using 10,000 simulations from the posterior GAM distributions (details in **Supplementary Methods**). Incidence is presented from 24-October-2022 to 12-February-2023 (weeks 3-18 of the respiratory pilot) as deconvolution tail estimates are highly uncertain. The infection duration was modelled using a Weibull distribution approximating ILI duration for ‘Flusurvey’ respondents^30^ (shape and scale parameters to match reported median (9 days) and IQR (reported=6-15 days, approximated=5-15 days). Due to insufficient data on appropriate distributions for influenza and RSV in community settings, other infection duration distributions were considered in sensitivity analyses (**Supplementary Table S1**).^31-35^

The probability of testing SARS-CoV-2-positive by age, conditional on reporting specific symptoms, was estimated for the full CIS sample using logistic GAMs. Similar models in the respiratory pilot expanded the outcome to testing positive for influenza A/B, RSV, or SARS-CoV-2, versus no virus identified, using multinomial GAMs (assigning 12 respiratory pilot samples positive for two viruses to the virus with the lowest cycle threshold (Ct) value). Both models included smooths for age and, for SARS-CoV-2 positivity in the larger sample, also calendar time, making predictions at 15-December-2022 to illustrate the contribution of SARS-CoV-2 to reported symptoms when all three pathogens’ positivity was relatively high. We used negative binomial GAMs to estimate the percentage self-reporting ILI and other symptoms by age amongst those testing positive or negative for SARS-CoV-2 in the full sample, and testing positive for influenza A/B and RSV in the respiratory pilot, averaged across the study period.

The effect of self-reported influenza vaccination on influenza A/B, RSV and SARS-CoV-2 positivity was estimated using logistic GAMs, controlling for demographics (age, sex, household size (1, 2, 3+), ethnicity (white versus non-white due to small numbers), ever worked in patient-facing healthcare, ever reported long-term health conditions, SARS-CoV-2 vaccination and prior SARS-CoV-2 infection (details in **Supplementary Methods**). All models included smooths for calendar time, age, days since most recent SARS-CoV-2 vaccination, and days since start of most recent SARS-CoV-2 infection (the last two truncated at 365 days (reference category), also with binary variables for unvaccinated or non-infected versus ≥365 days). Influenza vaccination was self-reported (“Have you received a flu vaccination since the last assessment” Yes/No/Missing). As the vaccination date was not elicited, participants were considered vaccinated if they had reported an influenza vaccination at a strictly prior assessment or at the current assessment if the prior assessment was >45 days ago. Very few participants (<3%) reported influenza vaccination in 22/23 only (**Table 1**), so these were categorised in models with “Both 22/23 and 21/22”.

**Table 1.**
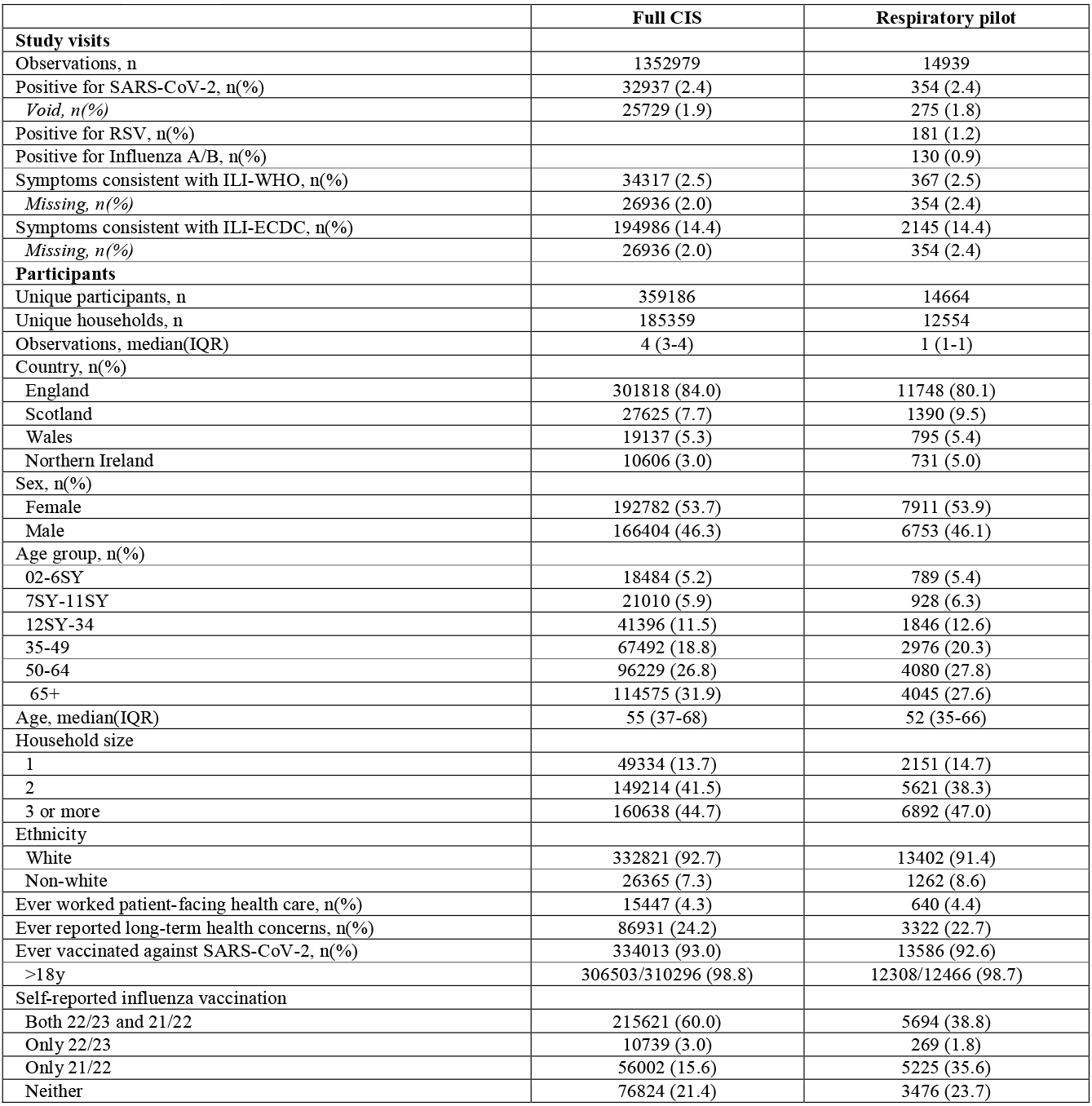
Study population characteristics.

|  | Full CIS | Respiratory pilot |
| --- | --- | --- |
| **Study visits** |  |  |
| Observations, n | 1352979 | 14939 |
| Positive for SARS-CoV-2, n(%) | 32937 (2.4) | 354 (2.4) |
| *Void, n(%)* | 25729 (1.9) | 275 (1.8) |
| Positive for RSV, n(%) |  | 181 (1.2) |
| Positive for Influenza A/B, n(%) |  | 130 (0.9) |
| Symptoms consistent with ILI-WHO, n(%) | 34317 (2.5) | 367 (2.5) |
| *Missing, n(%)* | 26936 (2.0) | 354 (2.4) |
| Symptoms consistent with ILI-ECDC, n(%) | 194986 (14.4) | 2145 (14.4) |
| *Missing, n(%)* | 26936 (2.0) | 354 (2.4) |
| **Participants** |  |  |
| Unique participants, n | 359186 | 14664 |
| Unique households, n | 185359 | 12554 |
| Observations, median(IQR) | 4 (3-4) | 1 (1-1) |
| Country, n(%) |  |  |
| England | 301818 (84.0) | 11748 (80.1) |
| Scotland | 27625 (7.7) | 1390 (9.5) |
| Wales | 19137 (5.3) | 795 (5.4) |
| Northern Ireland | 10606 (3.0) | 731 (5.0) |
| Sex, n(%) |  |  |
| Female | 192782 (53.7) | 7911 (53.9) |
| Male | 166404 (46.3) | 6753 (46.1) |
| Age group, n(%) |  |  |
| 02-6SY | 18484 (5.2) | 789 (5.4) |
| 7SY-11SY | 21010 (5.9) | 928 (6.3) |
| 12SY-34 | 41396 (11.5) | 1846 (12.6) |
| 35-49 | 67492 (18.8) | 2976 (20.3) |
| 50-64 | 96229 (26.8) | 4080 (27.8) |
| 65+ | 114575 (31.9) | 4045 (27.6) |
| Age, median(IQR) | 55 (37-68) | 52 (35-66) |
| Household size |  |  |
| 1 | 49334 (13.7) | 2151 (14.7) |
| 2 | 149214 (41.5) | 5621 (38.3) |
| 3 or more | 160638 (44.7) | 6892 (47.0) |
| Ethnicity |  |  |
| White | 332821 (92.7) | 13402 (91.4) |
| Non-white | 26365 (7.3) | 1262 (8.6) |
| Ever worked patient-facing health care, n(%) | 15447 (4.3) | 640 (4.4) |
| Ever reported long-term health concerns, n(%) | 86931 (24.2) | 3322 (22.7) |
| Ever vaccinated against SARS-CoV-2, n(%) | 334013 (93.0) | 13586 (92.6) |
| >18y | 306503/310296 (98.8) | 12308/12466 (98.7) |
| Self-reported influenza vaccination |  |  |
| Both 22/23 and 21/22 | 215621 (60.0) | 5694 (38.8) |
| Only 22/23 | 10739 (3.0) | 269 (1.8) |
| Only 21/22 | 56002 (15.6) | 5225 (35.6) |
| Neither | 76824 (21.4) | 3476 (23.7) |

## Results

### SARS-CoV-2, influenza and RSV positivity and incidence

Between 10-October-2022 and 26-February-2023, the 20-week period when additional influenza/RSV surveillance was conducted and when BQ.1, CH.1.1 and XBB SARS-CoV-2 sub-lineages were co-circulating in the UK, 32,937 (2.4%) of 1,352,979 swab tests conducted at study assessments were SARS-CoV-2-positive (median (IQR) 4 (3-4) tests/participant, 359,186 unique participants) (**Table 1**). 14,939 (1.1%) randomly selected swabs from 14,664 unique participants were tested in the respiratory pilot, with similar SARS-CoV-2 positivity (n=354, 2.4%). RSV and influenza A/B positivity were lower, 1.2% (n=181) and 0.9% (n=130) respectively. There were 12 (0.08%) coinfections (4 SARS-CoV-2/influenza, 4 SARS-CoV-2/RSV, 4 influenza/RSV; 653 (4.4%) swabs positive for ≥1 of the three viruses). Of 130 influenza A/B positives, subtype could be identified from PCR for 87 (remainder too low viral load/high Ct to amplify); 80 (92.0%) were influenza A, 5 (5.7%) influenza B, and 2 (2.3%) both (from whole genome sequencing 8 H1N1, 40 H3N3, and 1 Victoria)^36^. Percentages reporting ILI over the study period were very similar between the respiratory pilot and full CIS sample, with only minor differences in sample demographics (**Table 1**).

SARS-CoV-2 positivity and reported ILI-WHO peaked in late December 2022, with similar trends across the pilot and full samples (**Figure 1)**. Both trends varied by age; SARS-CoV-2 positivity was higher for older versus younger participants, while reported ILI-WHO was higher amongst those in SY11 or younger. In the full sample, SARS-CoV-2 positivity was consistently higher than reported ILI-WHO amongst those ≥65y, and trends in reported ILI-WHO were similar between those testing SARS-CoV-2 negative and positive. RSV and influenza positivity peaked earlier in December 2022, and also varied by age over time, with higher rates in younger children, and earlier peaks in RSV than influenza and SARS-CoV-2, particularly for those ≥50y. Cycle threshold (Ct) values for SARS-CoV-2 followed positivity trends, being lower (i.e. higher viral load) when positivity was higher (**Figure S3**). ILI-ECDC was more common than ILI-WHO, but followed broadly similar trends over time; other symptoms were either approximately constant over time or had similar peaks around December 2022 **(Figure S4-S6)**.

**Figure 1.**
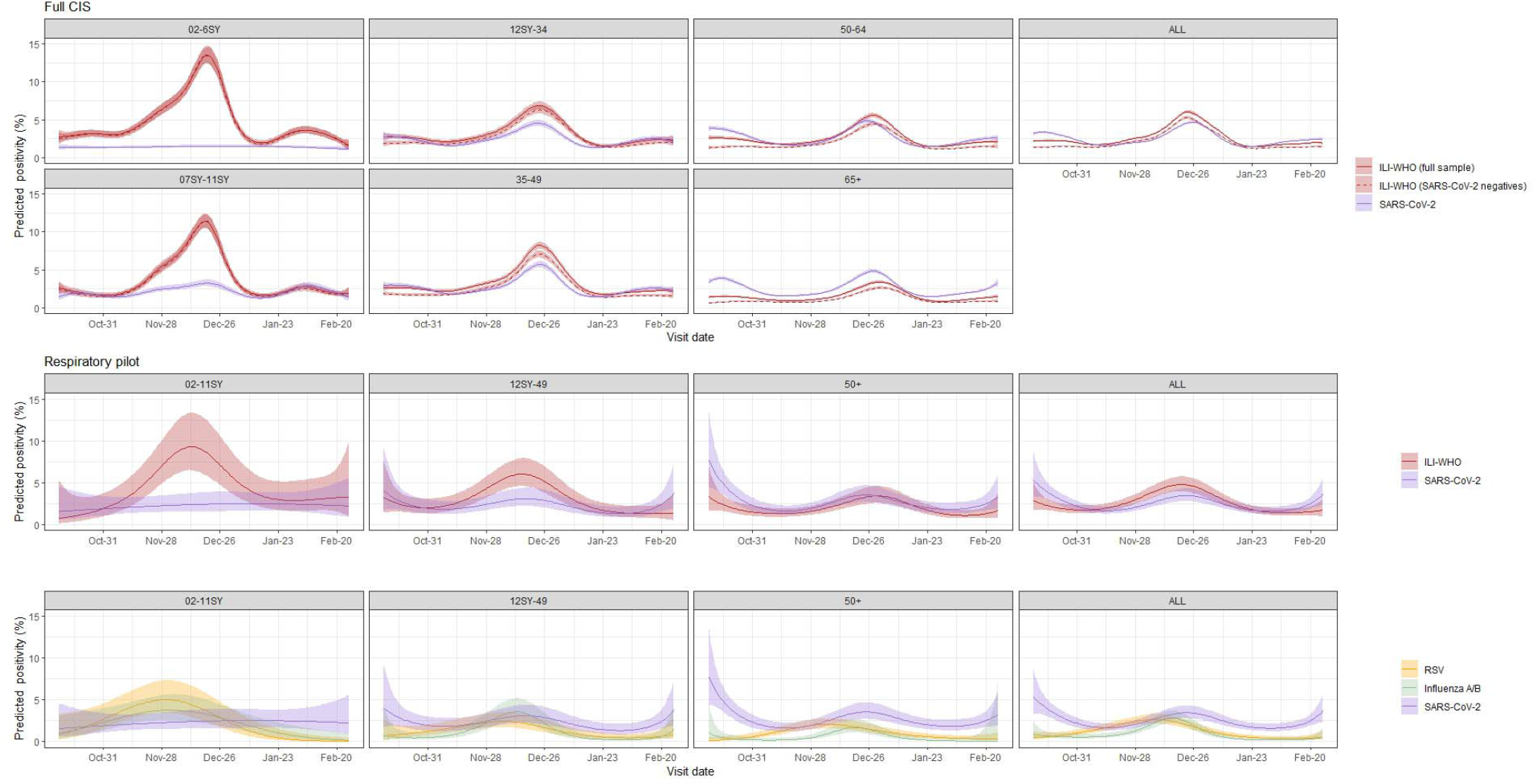
Percentage (95% CI) reporting ILI-WHO (full CIS and respiratory pilot) and test positivity for SARS-CoV-2 (full CIS and respiratory pilot), influenza A/B (respiratory pilot) and RSV (respiratory pilot) Note: SY=school-year. See supplementary material for raw daily percentages for the full CIS sample (**Figure S4**) and cumulative numbers positive for SARS-CoV-2, influenza A/B and RSV, and reporting ILI-WHO in the respiratory pilot (**Figure S5**).

Estimated incidence of SARS-CoV-2, influenza and RSV therefore also varied by age over time. In those 2-11SY, peak estimated daily incidence was higher and earlier for RSV (4.7/1000, 23-November-2022) and influenza (3.5/1000, 26-November-2022) than SARS-CoV-2 (2.8/1000, 17-December-2022), although overlapping credible intervals indicated considerable uncertainty **(Figure 2)**. For older age-groups, peak daily SARS-CoV-2 incidence (for 12SY-49 and 50+, 5.5 and 5.0/1000 on 18-December-2022 and 22-December-2022 respectively) was higher than RSV and influenza with similar shifts in timing. However, compared with younger children, peak daily RSV incidence was lower and later in older age-groups (2.2 and 1.9/1000 for 12SY-49 and 50+ respectively, both 30-November-2022) and peak daily influenza incidence also shifted later with increasing age (3.5 and 1.7/1000 on 6-December-2022 and 13-December-2022 for 12SY-49 and 50+ respectively). The choice of infection duration distribution did not alter the timing of the estimated peaks but influenced absolute incidence estimates **(Figure S7)**. Distributions with lower mean duration resulted in higher incidence, by approximately the inverse ratio of means (as expected from first-order approximations), so were ∼1.4 times higher using a distribution with mean 7.5^33^ versus 10.4 days (**Table S1**), although credible intervals overlapped for RSV and influenza.

**Figure 2.**
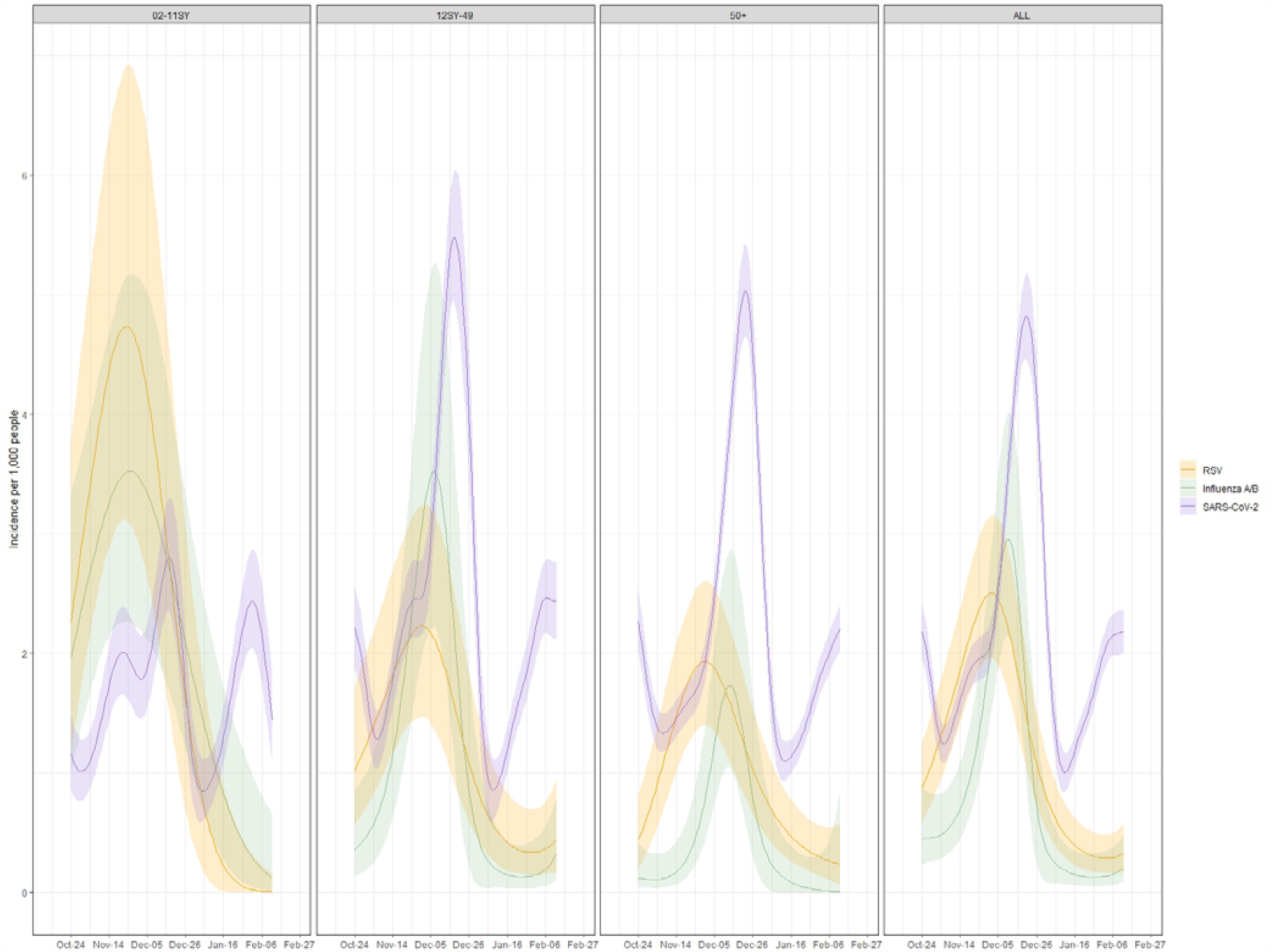
Estimated incidence (95%CI) of SARS-CoV-2 (full CIS), RSV (respiratory pilot), and influenza A/B (respiratory pilot) Note: Time frame covering October 24^th^ 2022 – February 13^th^ 2023. SY=school-year. Estimates based on a Weibull-ILI survival curve for infection duration. See supplementary material for further details on survival distributions (**Table S3, Figure S7**)

### Association between test positivity and self-reported symptoms

Considering age as a continuous variable (**Figure 3**), over 50% of SARS-CoV-2-positives aged 30-70y reported symptoms consistent with ILI-ECDC, compared to at most ∼25% in those 30-65y for ILI-WHO. ILI-ECDC symptoms were also more commonly reported than ILI-WHO amongst those testing positive for RSV or influenza, with ILI-WHO being particularly uncommon amongst RSV-positives, due to low rates of self-reported fever amongst RSV-positives across all ages. Cough and sore throat were amongst the most common symptoms for SARS-CoV-2-positives, with prevalence of cough >50% in those over ∼20y. However, in the youngest children, cough was almost as common in SARS-CoV-2-negatives as positives, consistent with multiple other causes. Sneezing, fatigue, and headache were other common symptoms amongst SARS-CoV-2-positives (**Figure S8**), with higher rates amongst middle-aged versus younger and older participants. As for SARS-CoV-2-positives, cough, sore throat, sneezing, fatigue and headache were amongst the most commonly reported symptoms for RSV- and influenza-positives, with broadly similar trends across age (**Figure 3, Figure S9**), including most symptoms being more commonly reported amongst middle-aged participants. Most symptoms were more commonly reported in influenza-than RSV-positives, wheezing being the main exception, being more commonly reported in older participants testing positive for RSV than influenza or SARS-CoV-2. However, absolute percentages reporting wheezing were lower than for other symptoms, and confidence intervals were wide.

**Figure 3.**
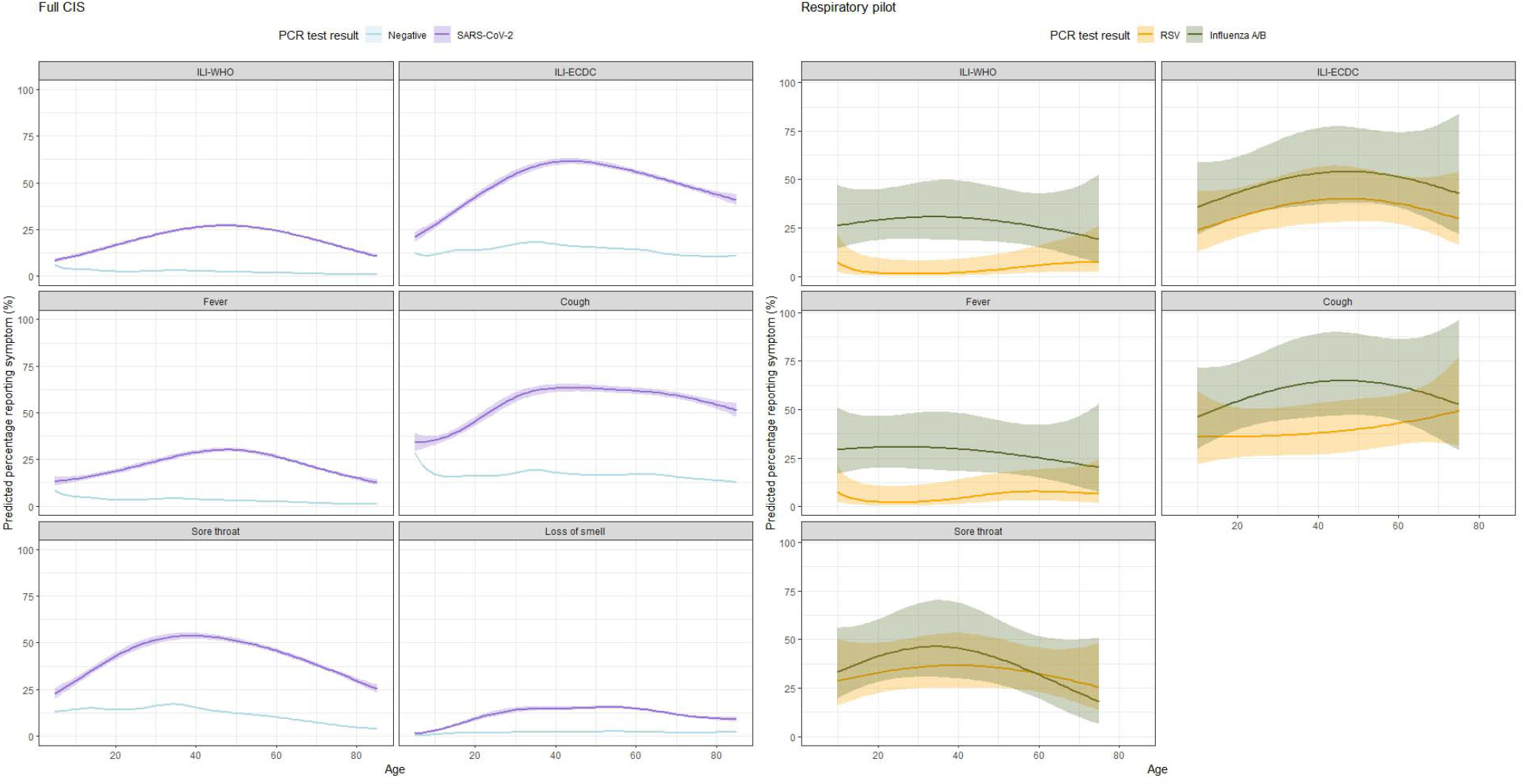
Prevalence of reported symptoms by SARS-CoV-2 test result (full CIS sample), and amongst those testing positive for RSV and influenza A/B (respiratory pilot) Note: see supplementary material, **Figures S8-S9** for the remaining symptoms. Predictions are averaged across time (no smooth for calendar time included in models). Respiratory pilot analysis excluded loss of smell due to small absolute number of participants reporting this symptom. Predictions were restricted to ages 10-75 years for the respiratory pilot due to the small absolute number outside this range (approximate 5^th^ – 95^th^ percentiles), and 5-85 years for the full CIS (approximate 1^st^ - 99^th^ percentiles).

Nevertheless, whether symptoms were defined by either ILI definition or individually, most (>65%) symptomatic (community-based) participants were not positive for SARS-CoV-2, influenza A/B, or RSV (**Figure 4, Figures S10-S11)**. The predicted probability of testing SARS-CoV-2-positive given specific symptoms generally increased with age, and was higher for ILI-WHO than ILI-ECDC. This appeared to be driven by higher probabilities of SARS-CoV-2 amongst participants reporting fever, the individual symptom with the largest percentage of confirmed viral cases in the full and respiratory pilot samples (**Figure 4)**. The respiratory pilot estimates suggested that, beyond SARS-CoV-2, RSV and influenza could only explain minor additional fractions of reported symptoms (**Figure 4, Figure S11**). Further, the probability of confirmed influenza infection tended to decrease with age amongst symptomatic participants, compared to the increasing trend for SARS-CoV-2, although uncertainty was relatively large (**Figure S13**).

**Figure 4.**
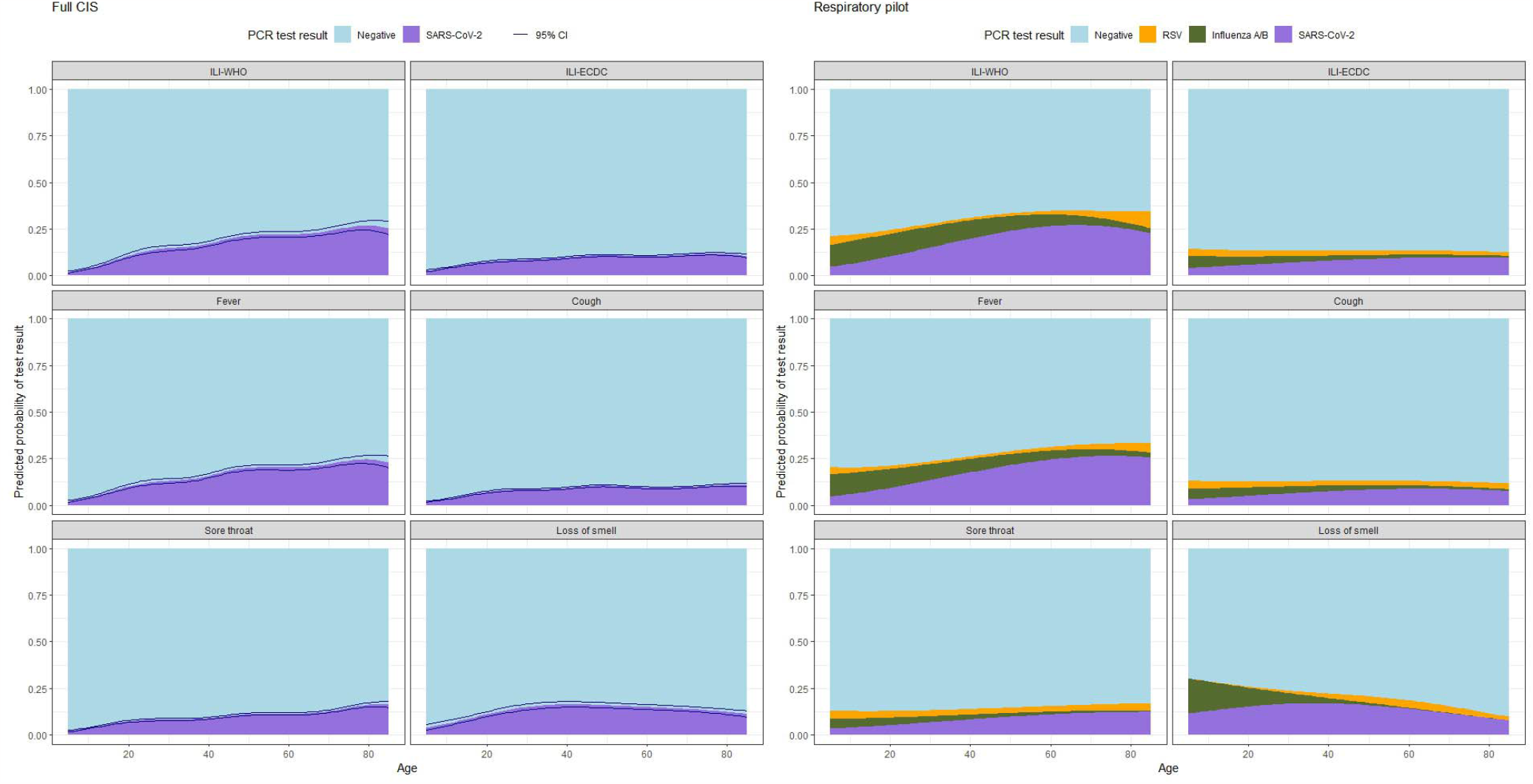
For participants reporting selected symptoms, predicted probabilities of a positive test result for SARS-CoV-2 on 15^th^ December 2022 (full CIS sample), and for SARS-CoV-2, influenza A/B or RSV (respiratory pilot sample), by age. Note: See supplementary material, **Figures S10-S11**, for the remaining symptoms. Predictions for the full CIS sample were made on 15^th^ December 2022 from models which adjusted for time, results for additional dates are shown in **Figure S12**. Predictions for the respiratory pilot are from a model not adjusted for time (given the limited sample size) and therefore represent an overall average over time. Predictions were made for ages 5-85 (approx. 1^st^-99^th^ percentiles).

### Influenza vaccination

In the respiratory pilot (winter 22/23), influenza A/B positivity was significantly lower for those reporting influenza vaccination both in the current 22/23 and prior 21/22 season versus not reporting influenza vaccination in either season (adjusted OR=0.55 (95% CI 0.32,0.95)); there was no evidence of association with influenza vaccination only in the past 21/22 season (aOR=0.81 (0.52,1.26), heterogeneity p=0.125) **(Table 2, Figure S14)**. Influenza A/B positivity was higher in those working in patient-facing healthcare (aOR=2.51 (1.31,4.79)). There was very weak evidence of interaction between vaccination status and age for influenza vaccination in the current and previous season (categorising as ≥ versus <18y heterogeneity p=0.541 and 0.113, respectively, **Table S2**). Including a continuous interaction with age **(Figure S15)**, the decreased risk associated with current and previous vaccination was greatest amongst young children and older adults. There was no evidence of association between influenza vaccination and RSV positivity or between prior SARS-CoV-2 infection or vaccination and influenza A/B or RSV positivity **(Table 2, Figure S14, Figure S16)**. Interestingly, in the much larger full sample, influenza vaccination in the current and prior season was associated with a slightly elevated risk of SARS-CoV-2-positivity (aOR=1.10 (1.05,1.14)), and similarly only in the prior season (OR=1.09 (1.05,1.13)), heterogeneity p=0.688), consistent with competing risks between SARS-CoV-2 and influenza or influenza vaccination targeting those most vulnerable to respiratory infection. We found evidence of waning protection against SARS-CoV-2 positivity over time from previous SARS-CoV-2 vaccination, and from previous SARS-CoV-2 infection after the initial period of PCR positivity **(Figure S17)**, with residual small additional risk in those never identified as infected (aOR=1.10 (1.06,1.14) versus last infected >365 days previously) and slightly lower risk in those not reporting prior SARS-CoV-2 vaccination (aOR=0.81 (0.75,0.88) versus last vaccinated >365 days previously).

**Table 2.** Model estimates for influenza vaccination.

|  | Influenza A/B<br>Respiratory pilot |  | RSV<br>Respiratory pilot |  | SARS-CoV-2<br>Full CIS |  |
| --- | --- | --- | --- | --- | --- | --- |
|  | *OR (95% CI)* | *P-value* | *OR (95% CI)* | *P-value* | *OR (95% CI)* | *P-value* |
| **Flu vaccination 21/22 vs. Neither** | 0.81 (0.52, 1.26) | 0.359 | 1.22 (0.83, 1.80) | 0.307 | 1.09 (1.05, 1.13) | <0.001 |
| **Flu vaccination both 21/22 and 22/23 vs. Neither** | 0.55 (0.32, 0.95) | 0.032 | 0.75 (0.46, 1.21) | 0.236 | 1.10 (1.05, 1.14) | <0.001 |
| **No SARS-CoV-2 vaccination** | 1.05 (0.49, 2.23) | 0.900 | 1.45 (0.75, 2.81) | 0.268 | 0.81 (0.75, 0.88) | <0.001 |
| **No prior SARS-CoV-2 infection** | 0.91 (0.53, 1.56) | 0.732 | 0.85 (0.54, 1.35) | 0.498 | 1.10 (1.06, 1.14) | <0.001 |
| **Upcoming SARS-CoV-2 vaccination in next 21 days** | - |  | - |  | 0.44 (0.40, 0.48) | <0.001 |
| **Female vs. Male** | 0.87 (0.61, 1.23) | 0.432 | 0.67 (0.50, 0.90) | 0.009 | 0.94 (0.93, 0.98) | <0.001 |
| **Ethnicity non-white vs. Ethnicity white** | 1.21 (0.72, 2.03) | 0.472 | 0.84 (0.49, 1.44) | 0.527 | 0.88 (0.84, 0.92) | <0.001 |
| **Household size 2 vs. Household size 1** | 1.66 (0.80, 3.44) | 0.173 | 1.11 (0.68, 1.83) | 0.671 | 1.11 (1.07, 1.14) | <0.001 |
| **Household size 3+ vs. Household size 1** | 1.37 (0.64, 2.91) | 0.419 | 0.86 (0.50, 1.47) | 0.584 | 1.13 (1.09, 1.18) | <0.001 |
| **Ever worked in patient-facing health care** | 2.51 (1.31, 4.79) | 0.005 | 1.01 (0.47, 2.17) | 0.986 | 0.94 (0.88, 0.99) | 0.029 |
| **Ever reported long-term health concerns** | 1.15 (0.72, 1.83) | 0.569 | 1.08 (0.74, 1.57) | 0.696 | 1.02 (1.00, 1.05) | <0.001 |

NOTE: All models include smooths for age, calendar time, days since most recent SARS-CoV-2 vaccination (truncated at 365 days), and days since start of most recent SARS-CoV-2 infection episode (truncated at 365 days). Estimated smooths can be seen in figures **S14, S16-S17**. The SARS-CoV-2 model also controls for upcoming SARS-CoV-2 vaccination in the next 21 days, as individuals testing SARS-CoV-2 positive were asked to defer vaccination (reverse causality), leading to a low probability of vaccination amongst those with a very recent infection. SARS-CoV-2 vaccination coinciding with study visit dates was counted from the next study visit onwards, flu vaccination coinciding with study visit dates were counted from the next study visit onwards unless the participant had not had any study visits in the past 45 days, in which case vaccination counted from the current visit.

## Discussion

Estimates from the full ONS CIS and its respiratory pilot suggest that positivity and incidence of SARS-CoV-2, influenza, and RSV varied by age and time across the 22/23 winter season. Peak incidence rates appeared somewhat delayed with increasing age for each virus, but particularly for influenza, with peaks observed approximately two weeks later for those 50y+ versus children 11SY or below. RSV peaked before influenza, and then SARS-CoV-2 in each age-group, although peaks were relatively close. Increasing influenza cases amongst children could hence provide an early warning for older age-groups, consistent with the former being the key driver of influenza transmission,^37^ and supporting early timing of childrens’ vaccination programmes to reduce overall transmission. We also observed higher RSV positivity and incidence for those 2-11SY versus older age-groups, and lower influenza positivity/incidence for those ≥50y.

A large fraction of symptoms reported by participants could not be attributed to test positivity for SARS-CoV-2, influenza A/B or RSV. This highlights the role of other infections not included in this study in symptom trends, including rhinovirus, adenovirus, human metapneumovirus, and parainfluenza as identified in syndromic surveillance,^38^ plus bacterial causes.^39^ Given the high prevalence of background symptoms observed in SARS-CoV-2-negatives, the symptoms reported by test positives for SARS-CoV-2, influenza A/B or RSV may not necessarily be caused by these infections specifically. RSV-positives generally tended to report fewer symptoms than SARS-CoV-2- or influenza-positives, but symptomatology generally appeared more strongly influenced by age than aetiology. Cough, sore throat, sneezing, fatigue and headache were all among the most commonly reported symptoms for each of the three infections, suggesting that discriminating between SARS-CoV-2, influenza and RSV based on symptoms alone may prove challenging, with implications for antiviral treatment and testing. Overall, our findings highlight that in the community, the contributions of these three pathogens to overall symptomatology appear modest. While ILI-ECDC was more commonly reported than ILI-WHO across all ages for the three infections, only ∼15% of reported ILI-ECDC could be explained by test positivity for SARS-CoV-2, influenza A/B, or RSV. Careful consideration of background rates and age-specific dynamics are thus necessary when using self-reported symptoms from community cohorts as a surveillance method for respiratory illness, highlighting the potential benefits of more flexible ILI definitions.^7,15^ Our findings of higher rates of self-reported symptoms in middle-aged participants, broadly consistent across symptoms and the three infections studied, raises important questions regarding the role of age in infection susceptibility, illness natural history, reporting behaviour, and vulnerability to other symptom-inducing conditions.

Although we confirmed previous findings of high rates of cough in test-negatives,^14^ we also found evidence of particularly high rates in older RSV-positives. This was the only symptom that approached rates of 50% amongst RSV-positives and confirms prior findings of cough’s relevance to RSV discrimination.^12,40,41^ In contrast, fever was rarely reported amongst RSV-positives across all ages. Fever has previously been identified as an important predictor of influenza,^14,16,42^ and we also found it was more commonly reported with influenza than SARS-CoV-2 or RSV for those <20y. Consequently, fever may have higher value for predicting influenza in children, yet it was also relatively common amongst SARS-CoV-2-positives. ILI-WHO and ILI-ECDC were similarly reported in SARS-CoV-2-positives and influenza-positives, indicating that the emergence of SARS-CoV-2 may complicate surveillance specifically targeting influenza. As previously suggested, ILI-WHO appears poorly suited to monitor RSV in the community^43-45^, due to its inclusion of fever.

Lower specificity and sensitivity of ILI definitions in our community sample compared to those presenting to clinical settings is perhaps unsurprising; however, one limitation is that the approximate monthly testing intervals in the full sample (from which the respiratory pilot was randomly selected) may also have affected the likelihood of symptom reporting, since questionnaires elicited symptoms in the last 7 days. For example, the design will have resulted in cases being identified at differing timepoints in their infection, so that those in a later stage of illness (or experiencing prolonged viral shedding) may appear asymptomatic at assessment although having experienced symptoms earlier in their infection, or may not test positive any longer despite still having symptoms. When positivity rates were low, Ct values supported a larger fraction of cases being identified late in infection (**Figure S3**). Another limitation is that we lacked information on the onset of individual symptoms, as all symptoms experienced within the past week were jointly reported. Further, the likelihood of reporting symptoms consistent with ILI is affected by other demographic factors including gender;^46^ we chose to focus on age as the main determinant of symptomatology, determinant of vaccination strategies and hence target of surveillance.

The main limitation is the smaller sample size in the respiratory pilot (which still tested ∼15,000 swabs), leading to greater uncertainty given the low event rates of RSV and influenza A/B. Although much smaller than the sample tested for SARS-CoV-2, this was still one of the larger community studies to date. Although broadly representative, non-white ethnicities and younger ages remained slightly under-represented, and SARS-CoV-2 vaccination slightly over-represented (although this has been shown to have short-lived effects on infection). Similarly, the limited data on infection duration distributions for RSV and influenza meant incidence estimates were approximate, although the choice of distribution affected absolute levels rather than relative rates or timing of peaks. Furthermore, the 22/23 winter season may not yet equate to steady-state post-pandemic mixing patterns in older adults.^47^ Influenza A and B were not differentiated in the multiplex assay, although the vast majority were A on further PCR (only successful in 67%), and we did not consider the impact of SARS-CoV-2 variant on symptomatology. During the study period, BQ.1, CH1.1 and XBB sub-lineages were co-circulating, and the high Ct values (low viral load) of many SARS-CoV-2-positives precluded universal sequencing to identify variants. Nevertheless, prior studies suggest that any symptom differences between influenza A and B are due to age and other risk factors.^8,9^

We found that influenza vaccination in both the current (22/23) and prior (21/22) season was associated with ∼45% protection against influenza test positivity in this general community sample, with no evidence of effect of vaccination in the prior season (21/22) only (point estimate ∼20% reduction). Numbers were too few to robustly assess the impact of vaccination in 22/23 only, although a recent test-negative case-control study suggested this group could have slightly greater benefit.^48^ Similarity in influenza strains included in the vaccine across the two seasons means that prior vaccination might have conferred some protection in the 22/23 season.^49^ Alternatively behavioural patterns or other factors differentiating those choosing vaccination could affect positivity. While Live Attenuated Influenza Vaccine (LAIV) could lead to vaccination-induced test positivity in children <18y, our estimates of protection were similar restricting to ≥18y, suggesting that effects of vaccination can still be identified in relatively small community cohorts.

In conclusion, our findings highlight the complex relationship between trends in test positivity for RSV, influenza A/B, and SARS-CoV-2, which peaked successively over the 22/23 winter season but to different degrees in different age-groups, and self-reported symptoms. Symptom profiles varied more by age than aetiology, making distinguishing between SARS-CoV-2, influenza and RSV on symptoms alone challenging, and most reported symptoms could not be explained by these viruses. Our findings emphasize the value of community-level data in understanding symptomatology in cases beyond those presenting to healthcare services, and have implications for COVID-19 contingency planning, particular in regards to the percentages not reporting respiratory symptoms.

## Data Availability

De-identified study data are available for access by accredited researchers in the ONS Secure Research Service (SRS) for accredited research purposes under part 5, chapter 5 of the Digital Economy Act 2017. For further information about accreditation, contact or visit the SRS website.

## Acknowledgements

We wish to thank all the individuals who participated in the COVID-19 Infection Survey.

We are grateful for the support of all the COVID-19 Infection Survey team:

Office for National Statistics: Sir Ian Diamond, Emma Rourke, Ruth Studley, Nick Taylor, Tina Thomas, Fiona Dawe;

Office for National Statistics COVID Infection Survey Analysis and Operations teams in particular Dawid Pienaar, Joy Preece, Sarah Crofts, Lina Lloyd, Michelle Bowen, Daniel Ayoubkhani, Russell Black, Antonio Felton, Megan Crees, Joel Jones, Esther Sutherland;

University of Oxford, Nuffield Department of Medicine: Ann Sarah Walker, Derrick Crook, Philippa C Matthews, Tim Peto, Emma Pritchard, Nicole Stoesser, Karina-Doris Vihta, Jia Wei, Alison Howarth, Kevin K Chau, Lucas Martins Ferreira, Brian D Marsden, Wanwisa Dejnirattisai, Juthathip Mongkolsapaya, Sarah Hoosdally, Richard Cornall, David I Stuart, Gavin Screaton;

University of Oxford, Nuffield Department of Population Health: Koen Pouwels;

University of Oxford, Big Data Institute: David W Eyre, Katrina Lythgoe, David Bonsall, Tanya Golubchik, Helen Fryer;

University of Oxford, Radcliffe Department of Medicine: John Bell;

Oxford University Hospitals NHS Foundation Trust: Stuart Cox, Kevin Paddon, Tim James;

University of Manchester: Thomas House;

UK Health Security Agency: Julie Robotham, Paul Birrell;

Office for Health Improvement and Disparities: John Newton, IQVIA: Helena Jordan, Tim Sheppard, Graham Athey, Dan Moody, Leigh Curry, Pamela Brereton;

National Biocentre: Ian Jarvis, Anna Godsmark, George Morris, Bobby Mallick, Phil Eeles;

Glasgow Lighthouse Laboratory: Jodie Hay, Harper VanSteenhouse;

Berkshire and Surrey Pathology Services: Muhammad Ehsaan;

Eric Haduli, Hugh Boothe, Reggie Samuel;

Welsh Government: Sean White, Tim Evans, Lisa Bloemberg;

Scottish Government: Katie Allison, Anouska Pandya, Sophie Davis;

Public Health Scotland: David I Conway, Margaret MacLeod, Chris Cunningham.

## Funding

This study was funded by the Department of Health and Social Care and the UK Health Security Agency with in-kind support from the Welsh Government, the Department of Health on behalf of the Northern Ireland Government and the Scottish Government. ED, EP, K-DV, KBP, ASW, TEAP, NS, DE are supported by the National Institute for Health Research Health Protection Research Unit (NIHR HPRU) in Healthcare Associated Infections and Antimicrobial Resistance at the University of Oxford in partnership with Public Health England (PHE) (NIHR200915). ASW and TEAP are also supported by the NIHR Oxford Biomedical Research Centre. KBP is also supported by the Huo Family Foundation. ASW is an NIHR Senior Investigator. PCM is funded by Wellcome (intermediate fellowship, grant ref 110110/Z/15/Z) and holds an NIHR Oxford BRC Senior Fellowship award. DWE is supported by a Robertson Fellowship and an NIHR Oxford BRC Senior Fellowship. NS is an Oxford Martin Fellow and an NIHR Oxford BRC Senior Fellow. JB is funded by the Bayes4Health grant. The views expressed are those of the authors and not necessarily those of the National Health Service, NIHR, Department of Health, or UKHSA. This work contains statistical data from ONS which is Crown Copyright. The use of the ONS statistical data in this work does not imply the endorsement of the ONS in relation to the interpretation or analysis of the statistical data. This work uses research datasets which may not exactly reproduce National Statistics aggregates. The funder/sponsor did not have any role in the design and conduct of the study; collection, management, analysis, and interpretation of the data; preparation, review, or approval of the manuscript; and decision to submit the manuscript for publication. All authors had full access to all data analysis outputs (reports and tables) and take responsibility for their integrity and accuracy. For the purpose of Open Access, the author has applied a CC BY public copyright licence to any Author Accepted Manuscript version arising from this submission.

## Author Contributions

The COVID-19 Infection Survey was designed and planned by ASW, ID and KBP, and was conducted by ASW, RS, NT, TEAP, PCM, NS, DWE, and the COVID-19 Infection Survey Team. This specific analysis was originally designed by ASW and ED, but with ongoing input from all authors on results. ED conducted the statistical analysis of the survey data. ED and ASW drafted the manuscript and all authors contributed to interpretation of the data and results and revised the manuscript. All authors approved the final version of the manuscript.

## Competing Interests statement

DWE declares lecture fees from Gilead, outside the submitted work. PCM has received GSK funding support.

## Notes

### Author Declarations

The South Central Berkshire B Research Ethics Committee (20/SC/0195) gave ethical approval for this study.

