## Supplementary Material for "SARS-CoV-2, influenza A/B and respiratory syncytial virus positivity and association with influenza-like illness and self-reported symptoms, over the 2022/23 winter season in the UK: a longitudinal surveillance cohort"

**Supplementary Methods**

***Cohort enrolment and management***

The ONS COVID-19 Infection Survey (CIS) was a large household survey with longitudinal follow-up (ISRCTN21086382; [https://www.ndm.ox.ac.uk/covid-19/covid-19-infection-survey/protocol-and-information-sheets](about:blank)). Private households were randomly selected from address lists and previous surveys on a continuous basis for enrolment from 26 April 2020 through 31 January 2022 (when new recruitment was paused, although follow-up continued until 13 March 2023 when study assessments were paused; 65% of participants were enrolled before December 2020, the start of the Alpha wave, and 84% before May 2021, the start of the Delta wave). Following verbal agreement to participate, a study worker visited each selected household to take written informed consent for individuals aged 2y and over. For those aged 2-15y, consent was provided by their parents or carers; those 10–15y also provided written assent. At the first visit, participants were asked for consent for optional follow-up assessments every week for the next month and then monthly subsequently (>97% consented to monthly follow-up). The survey received ethical approval from the South Central Berkshire B Research Ethics Committee (20/SC/0195).

At each assessment, participants were asked about demographics, behaviours (including testing positive on swabs taken outside of the survey), work, and vaccination status. Combined nose and throat swabs were taken from all consenting household members for SARS-CoV-2 PCR testing. After 31 July 2022, study worker visits were discontinued and participants could opt-in to continuing to complete questionnaires online or by telephone, returning test kits by post or courier (crossover 11-31 July 2022). There was no evidence that the change in mode of data collection in July 2022 (before the current study) influenced SARS-CoV-2 positivity^1,2^. There was evidence that all symptoms were more commonly reported with remote data collection (the current study was conducted only including data from the remote collection phase)^3^. Data collection was officially paused on 13 March 2023.

Age was grouped according to school years in children and young people, to reflect differences in mixing relate more to school than to age, specifically up to and including primary and middle school (school year (SY) 6), secondary school (SY7-SY11) and college or older (SY12 to 34). School years run from 1 September to 31 August the following year; SY6 is children who have their 11^th^ birthday in a school year, SY7 their 12^th^ birthday, SY11 their 16^th^ birthday and SY12 their 17^th^ birthday.

### ***Vaccination and prior infection***

Self-reported vaccination data were obtained from participants at assessments, including vaccination type, number of doses, and vaccination dates. Data from participants in England and Wales were also linked to administrative records from the National Immunisation Management Service (NIMS) in England and equivalent in Wales. We used records from administrative data sources where available and otherwise from the survey, since linkage was periodic and administrative data sources do not contain information about vaccinations received abroad or in Northern Ireland and Scotland. In GAMs assessing associations between influenza vaccination and test positivity, participants were counted as SARS-CoV-2 vaccinated if their vaccination date was strictly before the date of the study assessment. Participants were considered to have a prior SARS-CoV-2 infection based on positive PCR swab tests in CIS, positive tests from the linked national testing programme data from England and Wales (any type, including lateral flow tests), and self-reported positive test results (any type).

### ***Laboratory testing***

Combined nose and throat swabs from the CIS were analysed at the UK’s national Lighthouse Laboratories at Milton Keynes (through February 2021 only) and Glasgow using identical methodology. PCR for three SARS-CoV-2 genes (N protein, S protein, and ORF1ab) was performed using the Thermo Fisher TaqPath RT-PCR COVID-19 kit, and analysed using UgenTec FastFinder 3.300.5, with an assay-specific algorithm and decision mechanism that allows conversion of amplification assay raw data from the ABI 7500 Fast into test results with minimal manual intervention. Positive samples are defined as having at least a single N and/or ORF1ab gene detected, and PCR traces exhibited an appropriate morphology. The S gene alone is not considered to be positive.

For the respiratory pilot, swabs (identified only by a barcode with no other identifying features and arriving daily from a consolidation centre pooling deliveries from the postal service and couriers across the whole country) were randomly selected on receipt at the Glasgow laboratory and sent to Berkshire and Surrey Pathology Services for multiplex testing. Viral ribonucleic acid (RNA) extraction was performed using the Amplitude™ platform, which includes the Tecan™ Fluent™ 1080 Automation Workstation and the KingFisher™ Presto Purification Systems. We utilized the Thermo Fisher kit with Catalog Number A49598 for the extraction process. Subsequently, we conducted multiplex PCR tests on the extracted RNA using the Thermo Fisher TaqPath™ COVID-19, Flu A/B, RSV Combo Kit (Catalog Number A49867). This kit is designed to detect three different targets: SARS-CoV-2 (N/S combined), influenza A/B (combined), and Respiratory Syncytial Virus A/B combined (RSV). In a 384 well PCR plate, we added 6µl of the PCR reaction mix using a multichannel pipette followed by transferring 14µl of the extracted RNA employing the Agilent Bravo Platform. PCR plate was sealed with MicroAmp™ Optical Adhesive Film, vortexed and centrifuged. The PCR plate was then run in the Applied Biosystems™ QuantStudio™ 5 Real-Time PCR Instrument. After completing the PCR, we analyzed the results using QuantStudio™ Design and Analysis Software version 2.5. A Ct value of ≤45 was considered positive for influenza A/B and RSV. A Ct value of ≤37 was considered positive for SARS-CoV-2. Influenza A/B positives (combined result) were sent to the University of Oxford for whole genome sequencing which was used to determine subtype.

***Statistical methods – details of incidence estimation***

Incidence was estimated using Richardson-Lucy-type deconvolution from daily estimates of test positivity and the distribution of infection duration using 10,000 simulations from the posterior GAM distributions from the three wider age groups and overall. Posterior simulation from the GAM models was conducted with a simple Metropolis-Hastings sampler, as implemented in the gam.mh function from the *mgcv* package, scaling the posterior covariance matrix by 0.4 when generating random walk proposals and retaining every second sample. The Metropolis-Hastings sampler was used as the standard Gaussian approximation of the posterior can be poor for log-link models in periods of low positivity, observed towards the end of the respiratory pilot in particular^4^. Due to insufficient data on appropriate distributions for influenza and RSV in community settings, other infection duration distributions were considered in sensitivity analyses (**Supplementary Table S1**). Two were based on SARS-CoV-2 pre-Alpha and pre-vaccination (ATACCC study),^6^ the first slightly modified to ensure convergence and the latter incorporating additional information from CIS regarding long PCR positivity in the distribution tail. Weibull distributions were also used to approximate infection duration for influenza (median 7 days) and RSV (median 8 days) from the literature^8-10^ (the latter similar to Omicron period estimates from the National Basketball Association).^11^

***Representativeness***

The most recent Census data available is from 2021 for England and Wales, but only 2011 from Scotland and Northern Ireland. Therefore the current best estimates of the UK population are based on population projections made by the Office for National Statistics^5^. Of the total estimated UK population of 64,663,793, 31,190,622 (48.8%) were male and 32,753,171 (51.2%) were female.

8,112,866 (12.6%), 3,880,522 (6.0%), 5,886,258 (9.1%), 8,901,150 (13.8%), 12,732,174 (19.7%), 16,315,106 (25.2%) and 8,835,719 (13.7%) were 2-11, 12-16, 17-24, 25-34, 35-49, 50-69, and 70+ years, respectively (different age categories to those used in this analysis which were based on influenza vaccination eligibility; other breakdowns not available). 55,513,974 (85.9%), 4,791,530 (7.4%), 2,150,975 (3.3%), 1,064,744 (1.6%) and 1,142,570 (1.8%) were White, Asian/Asian British, Black/African/Caribbean/Black British, Mixed/Multiple ethnic groups and Other ethnic groups respectively.

**Table S1. Infection duration distributions used in main (bold) and sensitivity analyses for incidence**

| **Survival distribution** | Reference | Mean (area under the curve) | Median (IQR) of distribution used | Median (IQR) in the literature |
| --- | --- | --- | --- | --- |
| ATACCC | Seran at al. (2022)^5^ | 18.4 | 17 (13-22) |  |
| ATACCC with CIS tail* | Blake, personal communication | 20.1 | 16 (11-25) |  |
| **ILI (Weibull)** | **Camacho et al. (2013)^6^** | **10.4** | **9 (5-15)** | **9 (6-15)** |
| RSV (Weibull)** | Vos et al. (2020)^7^ | 10.5 | 8 (4-15) | 8 (5-14) |
| Flu (Weibull) | Vos et al. (2020)^7^ | 7.5 | 7 (5-11) | 7 (5-10) |

* ATACCC distribution with longer tail to reflect small number of individuals with very long durations of PCR positivity from CIS

** Similar median and early duration of infection distribution to estimates from the Omicron period from the National Basketball Association.

Note: ILI (Weibull) used for main analyses and is shown in bold. Distributions presented graphically in **Figure S7**.


**Table S2. Influenza vaccination effects on Influenza A/B test positivity, differential effects for Adults/Children**

|  |  | **Influenza A/B  Respiratory pilot** | |
| --- | --- | --- | --- |
|  |  | *OR (95% CI)* | *P-value* |
| **Flu vaccination 21/22 vs. Neither** | Adults =>18y | 0.65 (0.37, 1.14) | 0.135 |
|  | Children <18y | 1.15 (0.56, 2.35) | 0.703 |
| **Flu vaccination both 21/22 and 22/23 vs. Neither** | Adults =>18y | 0.53 (0.28, 1.01) | 0.056 |
|  | Children < 18y | 0.57 (0.21, 1.57) | 0.278 |
| **No SARS-Cov-2 vaccination** |  | 1.04 (0.49, 2.21) | 0.915 |
| **No prior SARS-Cov-2 infection** |  | 0.92 (0.54, 1.59) | 0.774 |
| **Female vs. Male** |  | 0.88 (0.62, 1.25) | 0.468 |
| **Ethnicity non-white vs. Ethnicity white** |  | 1.22 (0.73, 2.05) | 0.453 |
| **Household size 2 vs. Household size 1** |  | 1.68 (0.81, 3.48) | 0.165 |
| **Household size 3+ vs. Household size 1** |  | 1.36 (0.64, 2.90) | 0.426 |
| **Ever worked in patient-facing health care** |  | 2.61 (1.35, 5.00) | 0.004 |
| **Ever reported long-term  health concerns** |  | 1.15 (0.72, 1.85) | 0.552 |

**Figure S1. Raw percentages reporting ILI-WHO (full sample and amongst SARS-CoV-2-negatives) and test positivity for SARS-CoV-2 in the full CIS sample**

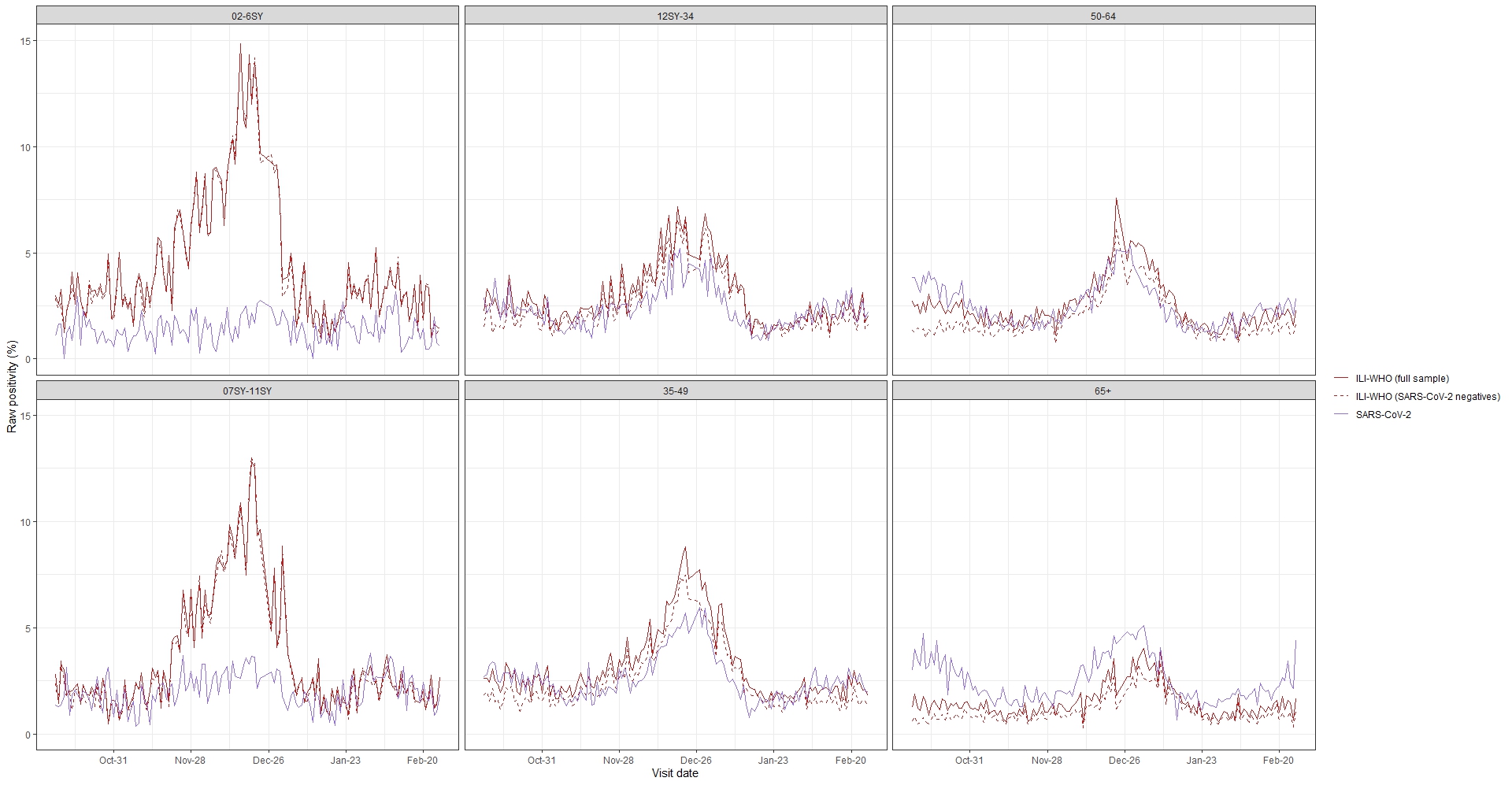

Note: Time frame covering October 10^th^ 2022 – February 26^th^ 2023, study assessments December 24-26^th^ and January 1^st^ excluded

**Figure S2.** **Cumulative number of study assessments reporting ILI-WHO, and cumulative numbers of positive PCR test results for SARS-CoV-2, influenza A/B, and RSV, in the respiratory pilot**

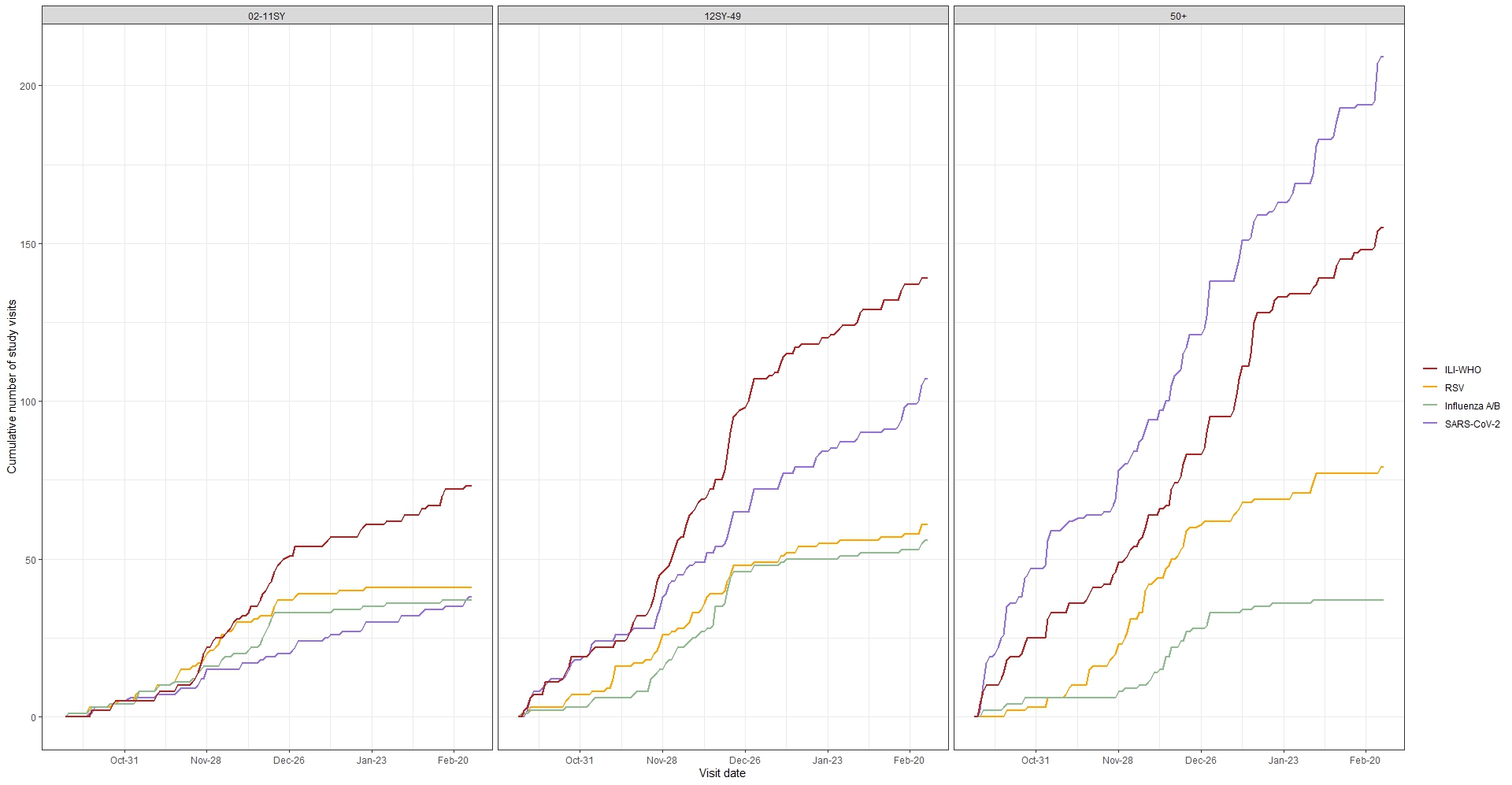

**Figure S3. Distribution of Ct values for positive PCR tests for influenza A/B and RSV (respiratory pilot), and for positive PCR tests for SARS-CoV-2 (full CIS) by month and week**
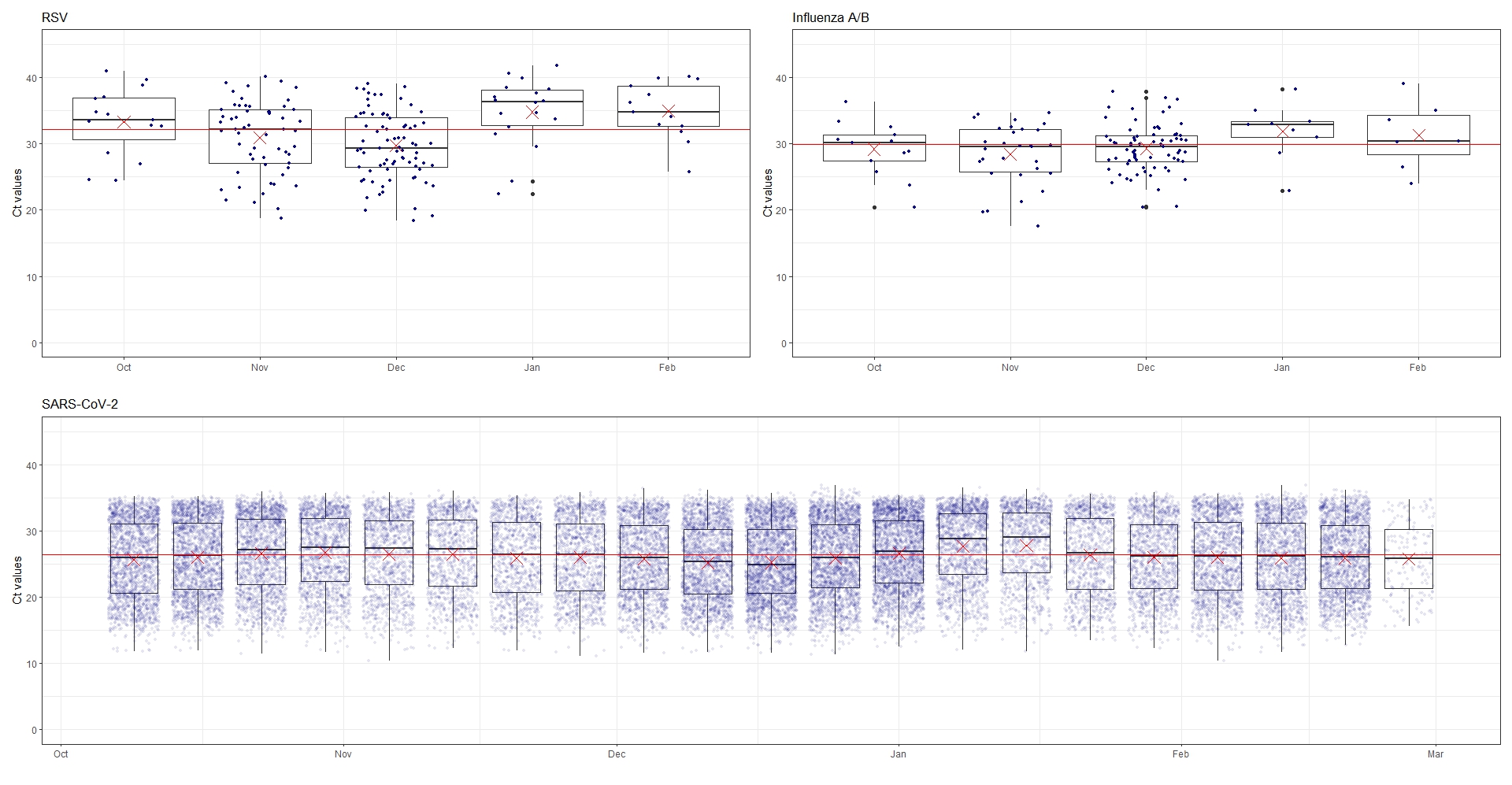

Note: Ct values for influenza A/B and RSV from the respiratory pilot are grouped by month of the study assessment, while Ct values for SARS-CoV-2 from the full CIS sample are grouped by week (week starting October 9^th^ 2022 – week starting February 20^th^ 2023). The red crosses indicate the monthly or weekly mean Ct value, while the red horizontal line indicates the overall median across the time period

**Figure S4. Estimated percentages reporting ILI-ECDC and respiratory symptoms in the respiratory pilot**

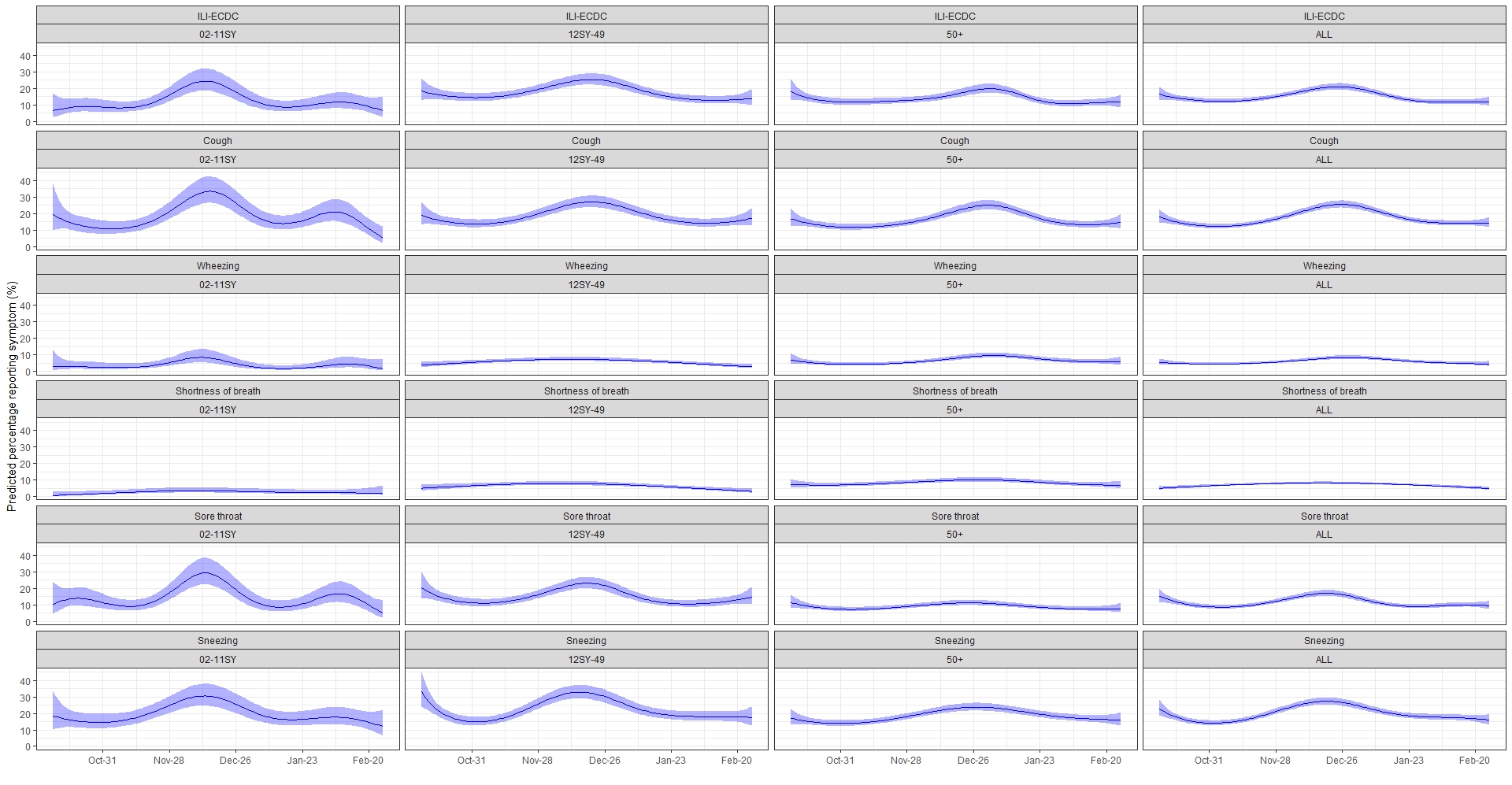

**Figure S5.** **Estimated percentages reporting systemic symptoms in the respiratory pilot**
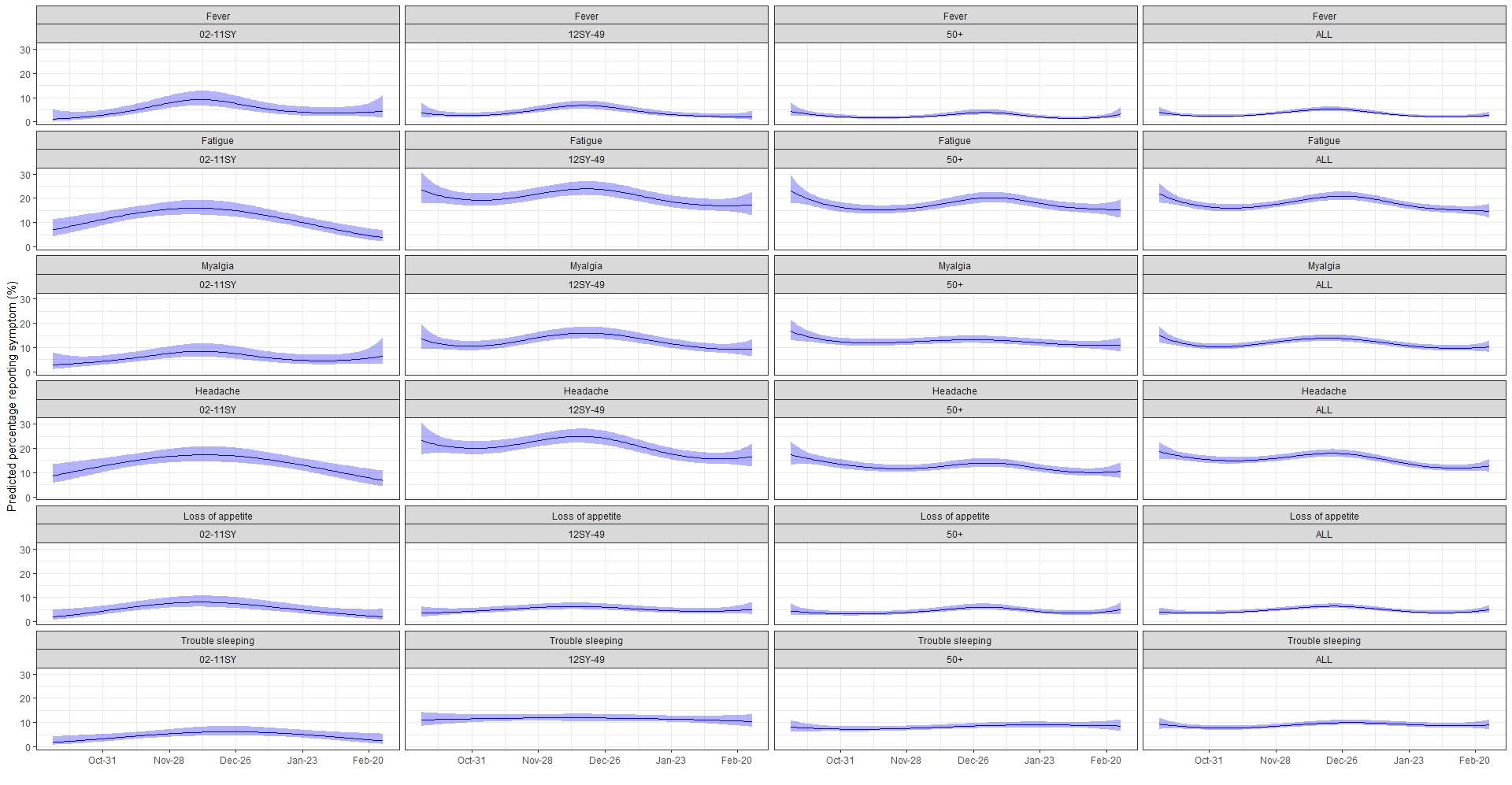

**Figure S6.** **Estimated percentages reporting loss of taste, loss of smell, and gastrointestinal symptoms in the respiratory pilot**
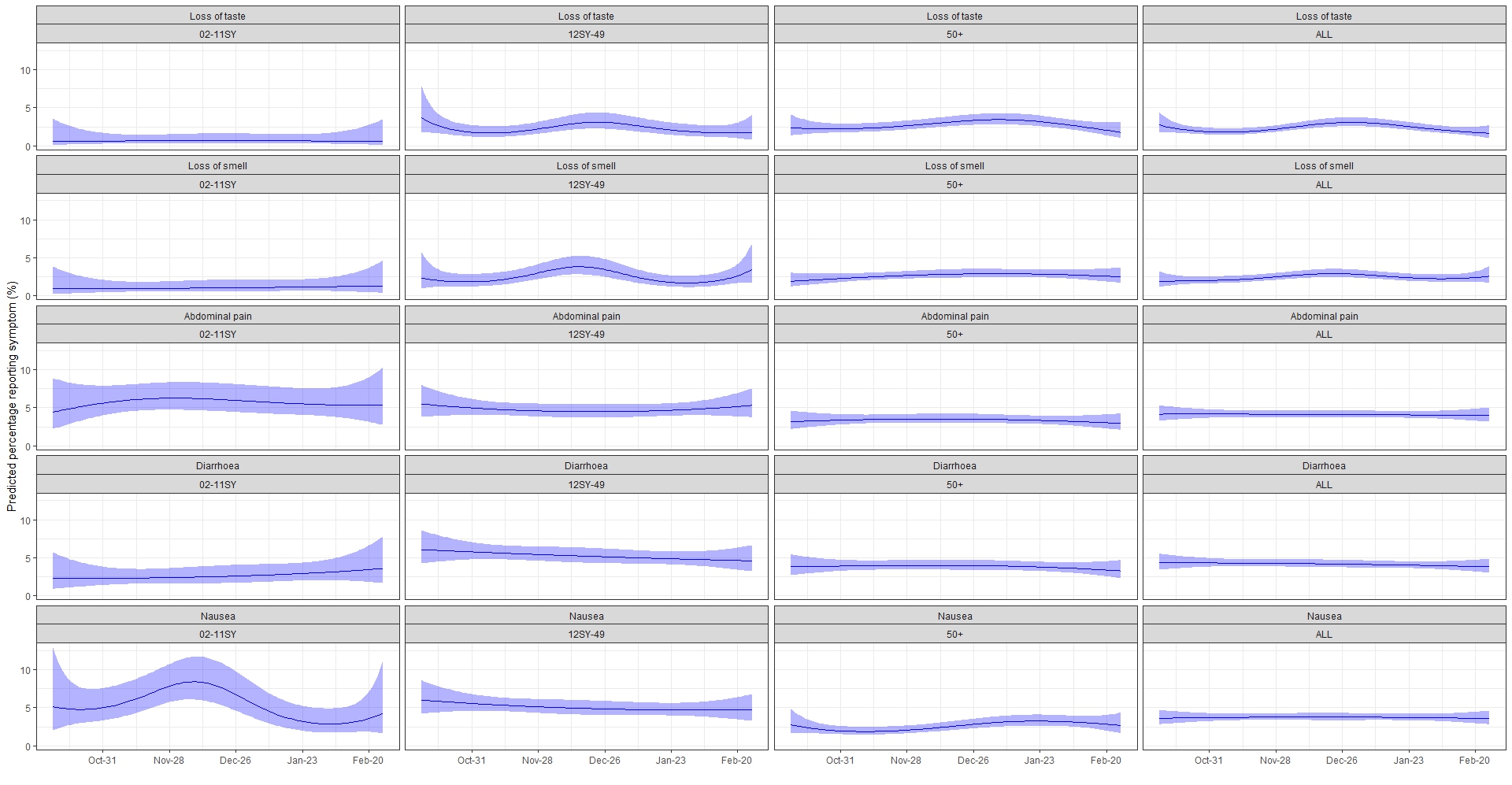

**Figure S7.** **Infection duration distributions (top left), and estimated incidence (total population) of SARS-CoV-2 (top right), RSV (bottom left), and influenza (bottom right) by infection duration distribution**
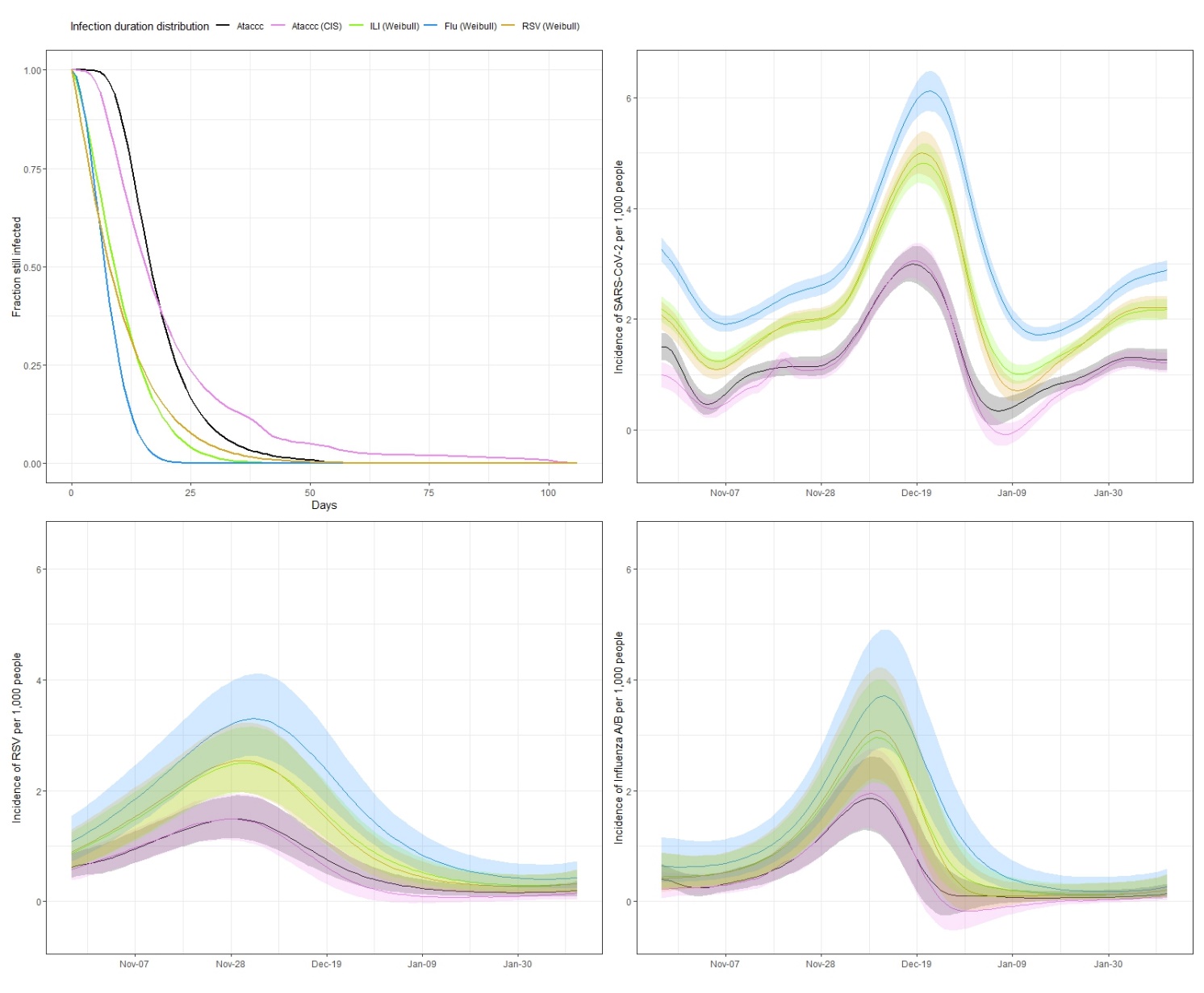

Note: ILI (Weibull) used in main analyses (**Figure 2**).

**Figure S8. Prevalence of reported symptoms over age by SARS-CoV-2 test result, Full CIS, remaining symptoms**
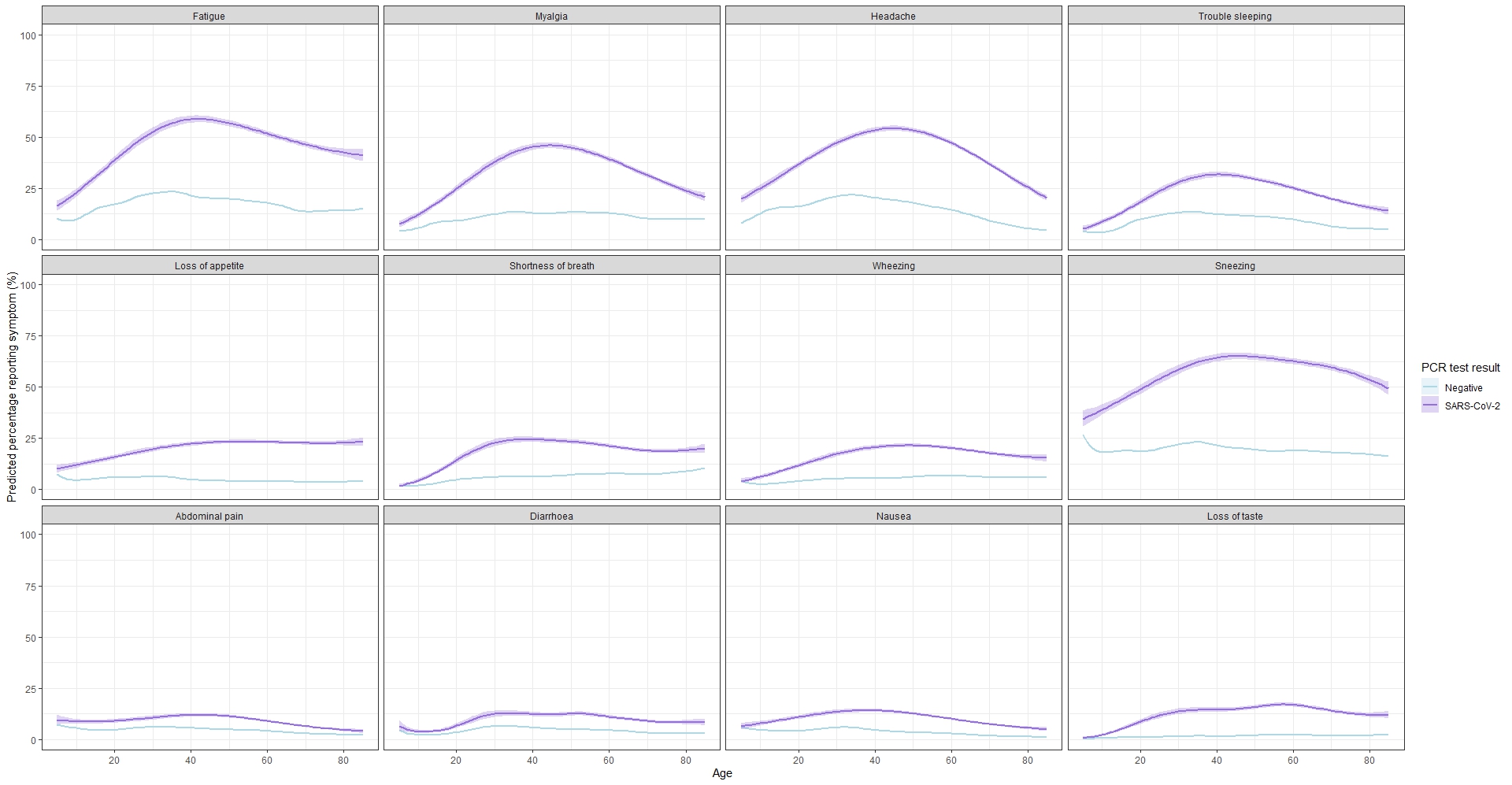

Note: Predictions restricted to ages 5-85 (approximate 1^st^ – 99^th^ percentiles)

**Figure S9. Prevalence of reported symptoms amongst those testing positive for RSV and influenza A/B, Respiratory pilot, remaining symptoms**
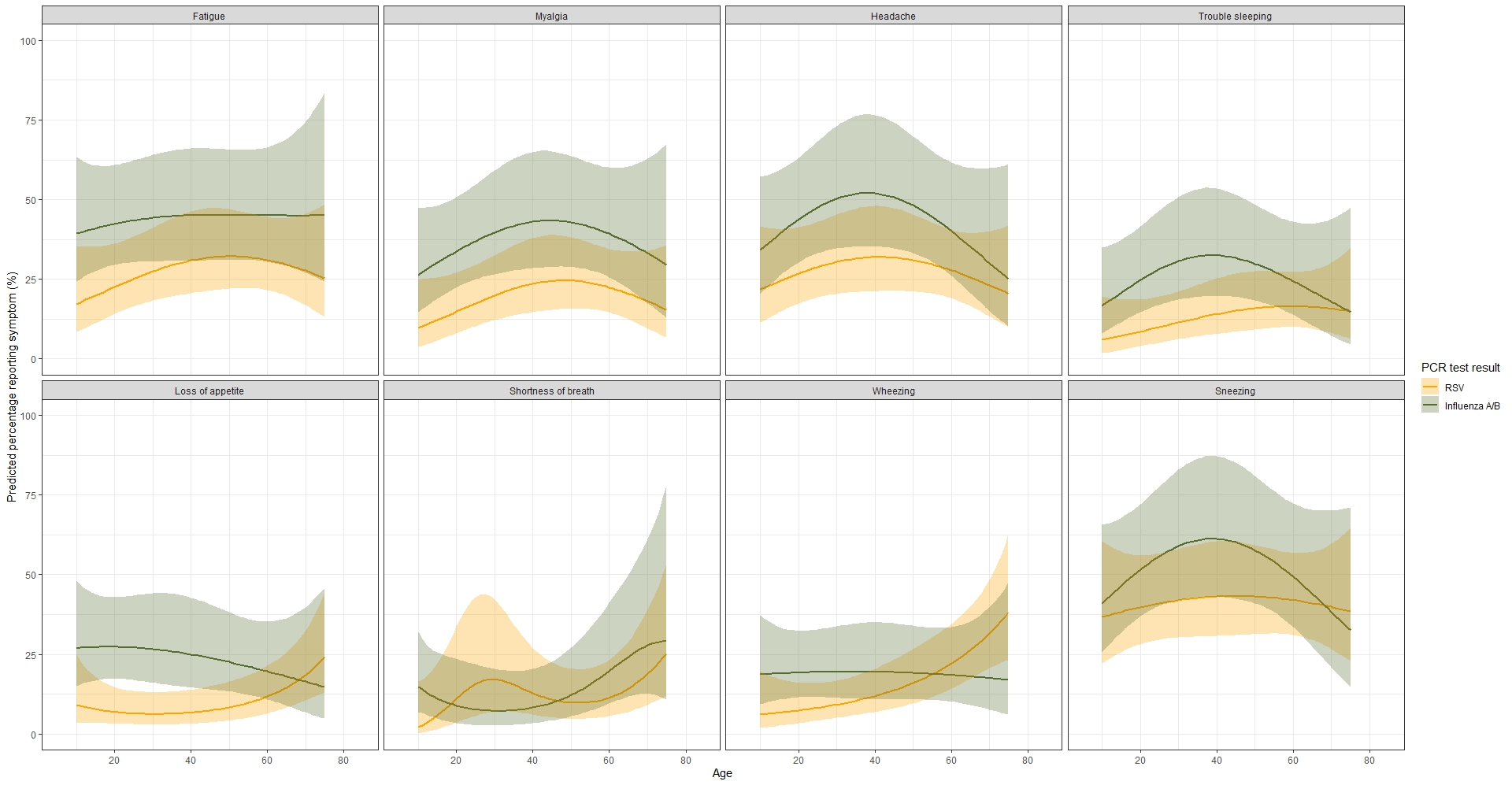

Note: Predictions restricted to ages 10-75 (approximate 5^th^ – 95^h^ percentiles). GI symptoms (abdominal pain, diarrhoea and nausea) and loss of taste/smell were not analysed for the respiratory pilot due to the small number of participants reporting these symptoms (less than 10% of Influenza A/B and RSV positives).

**Figure S10. Predicted probabilities of a positive test result for SARS-CoV-2 on December 15^th^, Full CIS, across age, remaining symptoms**

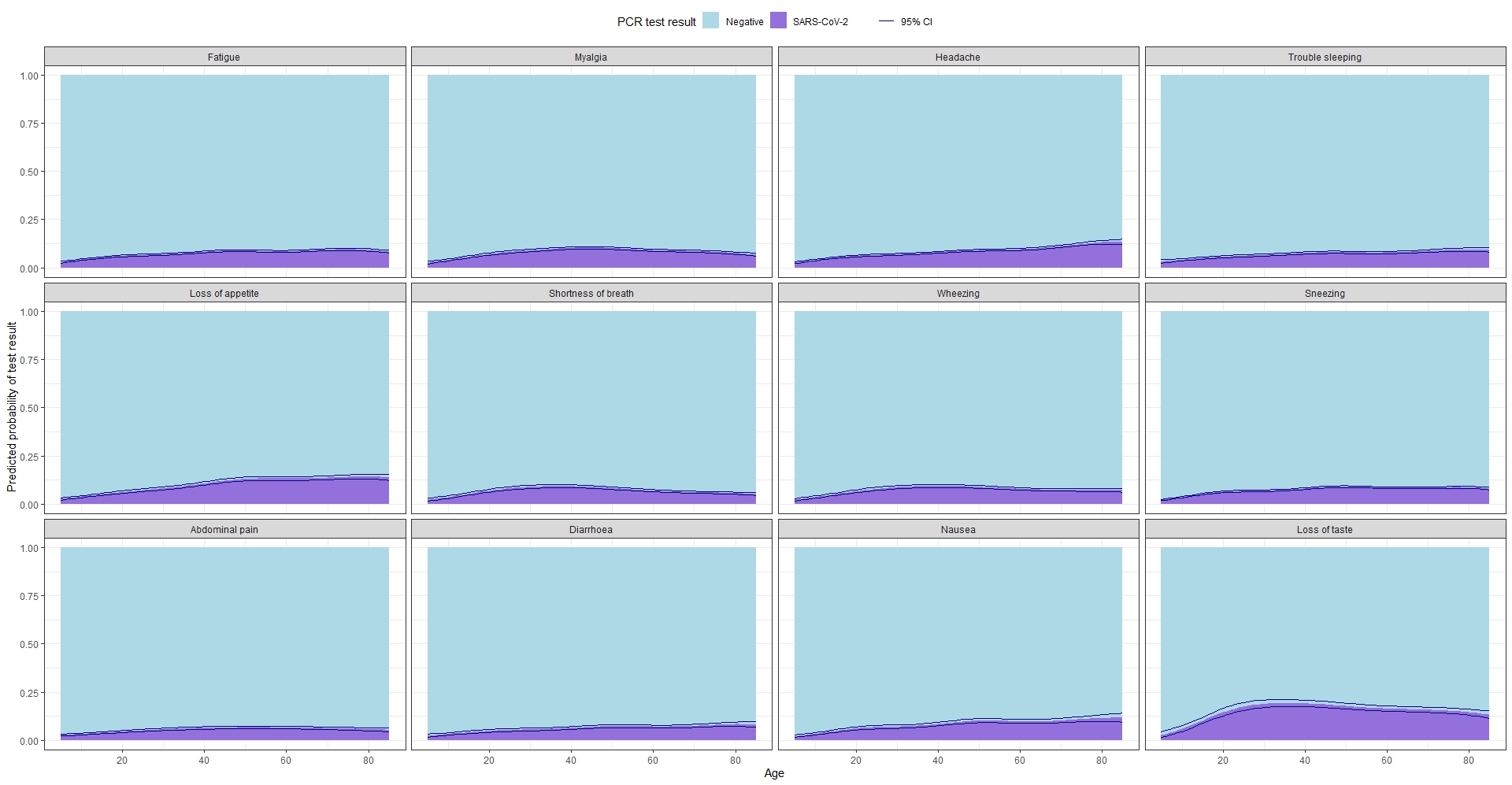

**Figure S11. Predicted probabilities of a positive test result for SARS-CoV-2, influenza A/B, or RSV, respiratory pilot, across age, remaining symptoms**

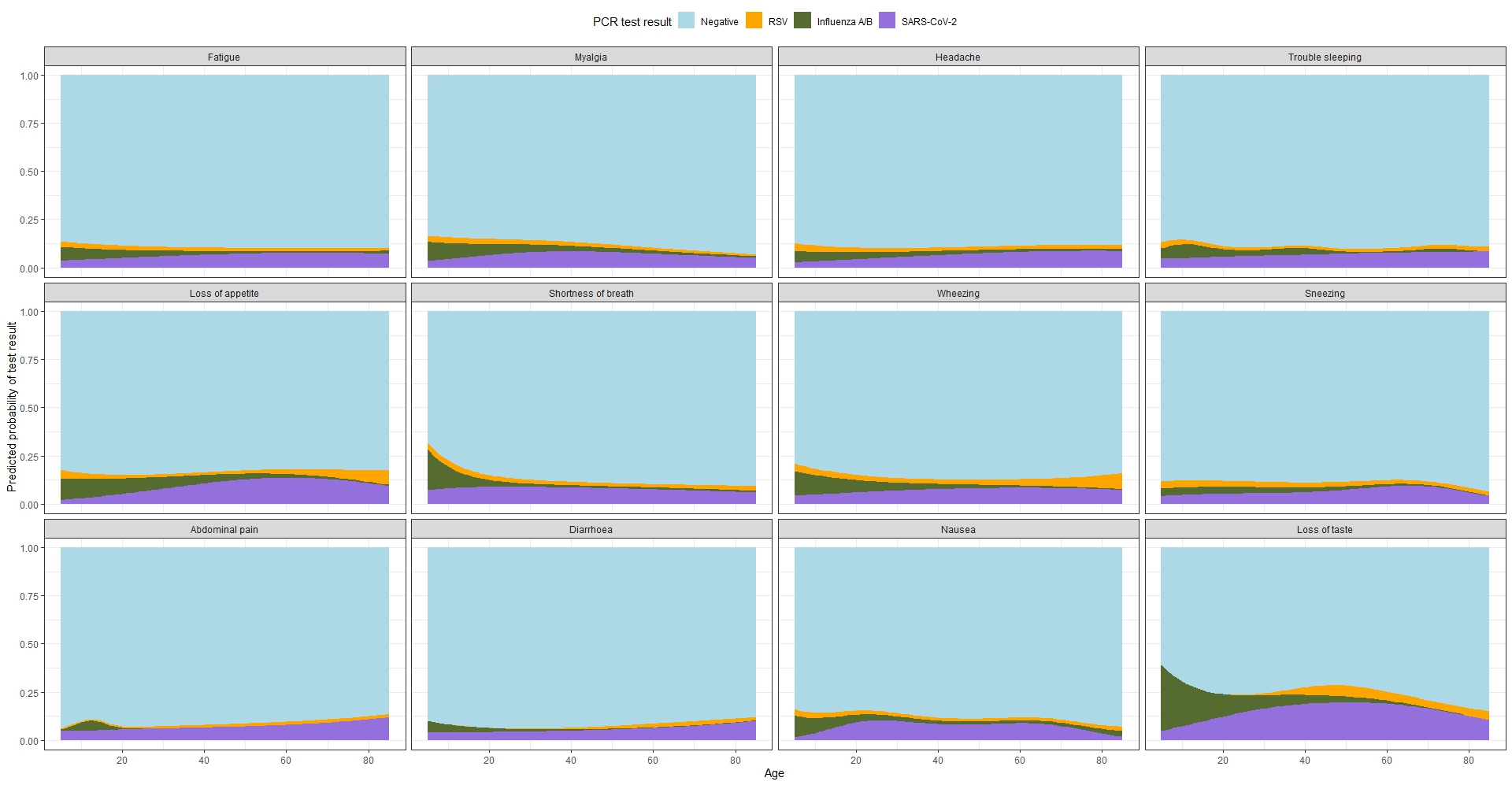

**Figure S12. Predicted probabilities of a positive test result for SARS-CoV-2on November 15^th^ 2022, December 15^th^ 2022, January 15^th^ 2023, and February 15^th^ 2023, across age, full CIS, selected symptoms**
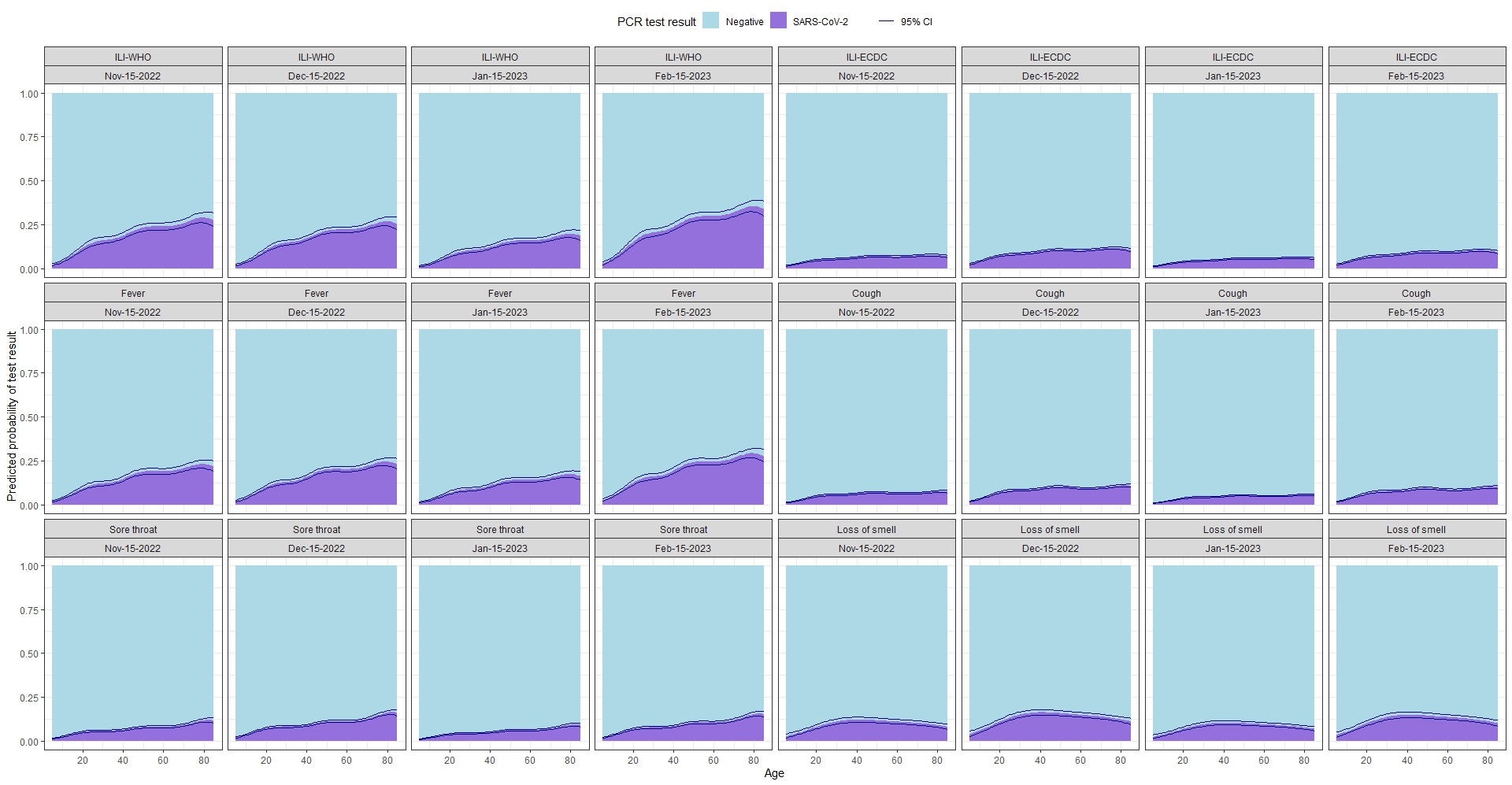

**Figure S13. Predicted probabilities of a positive test result for SARS-CoV-2, influenza A/B, or RSV, Respiratory pilot, across age, selected symptoms, with 95% CI**

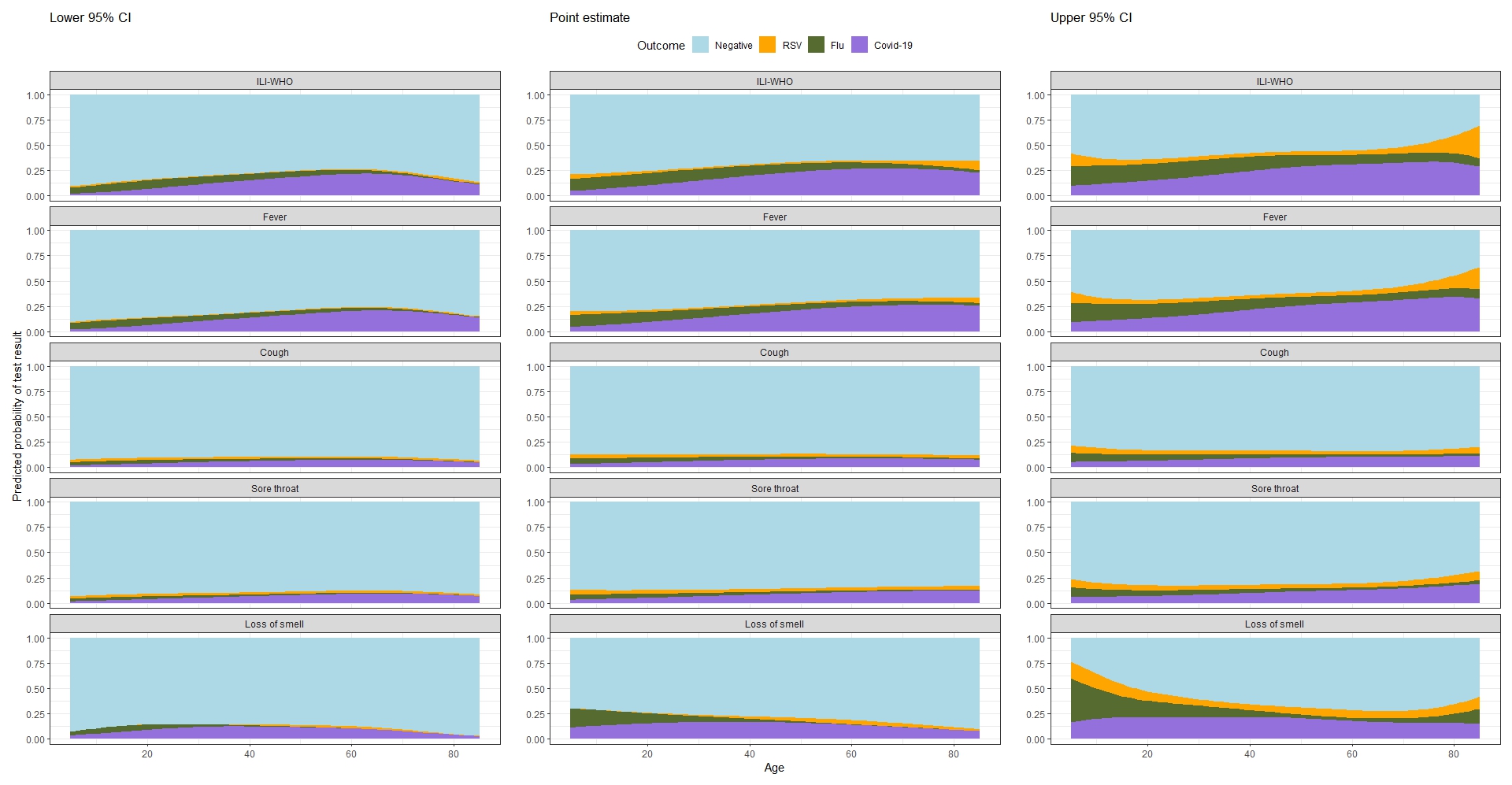

**Figure S14. Association between influenza A/B positivity and age, assessment date and days since most recent SARS-CoV-2 infection or vaccination in the respiratory pilot**
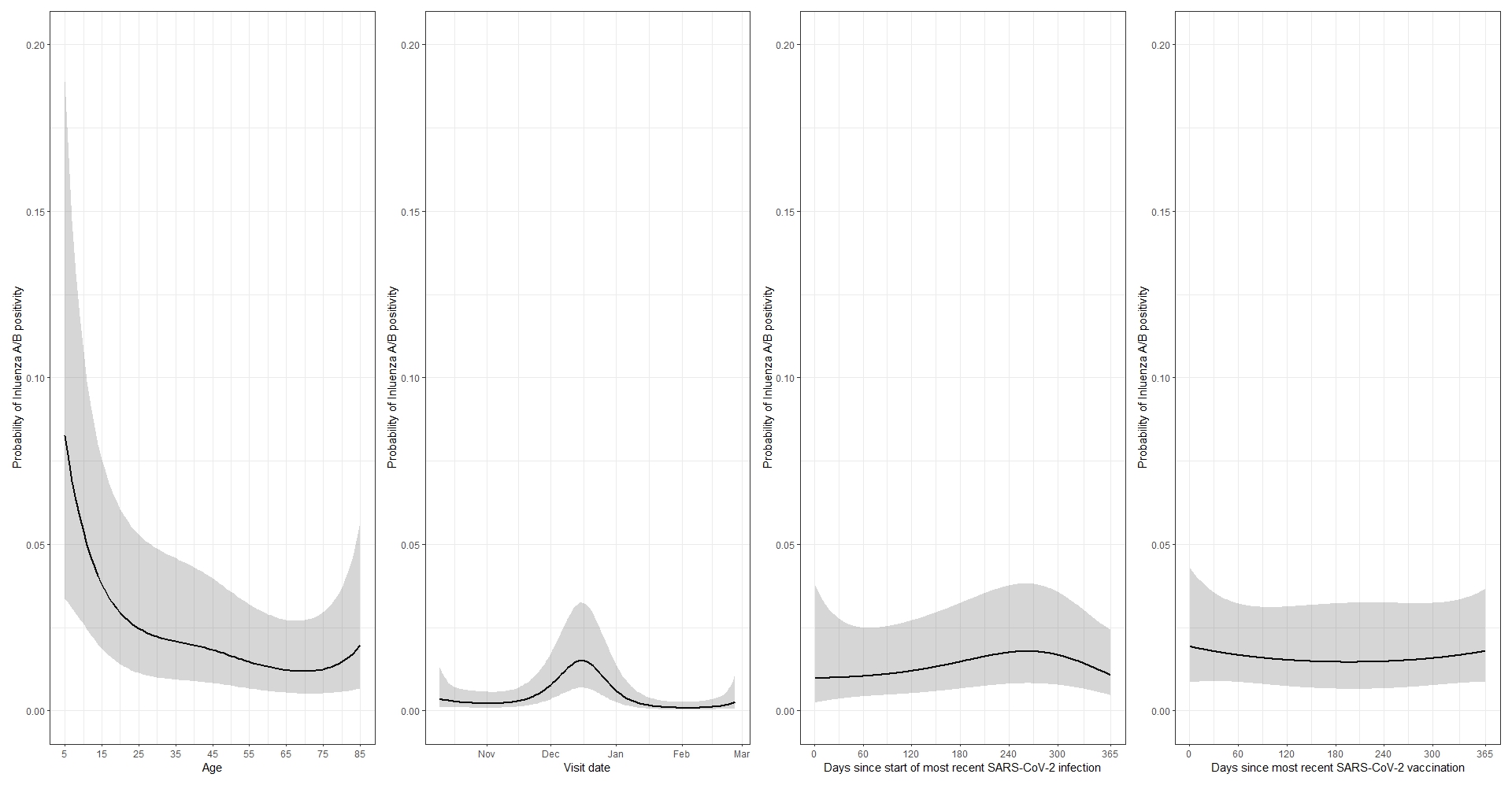

Note: Probabilities estimated at the following (approximate median for continuous variables): study day 70 (December 18^th^ 2022), Sex=Female, Ethnicity=White, Household size=3 or more, Ever reported long-term health concerns=No, Ever worked in patient-facing health care=No, Prior SARS-CoV-2 infection=Yes, Days since start of most recent SARS-CoV-2 infection = 259, Vaccinated against SARS-CoV-2=Yes, Days since most recent SARS-CoV-2 vaccination=165, Flu vaccination=Both 21/22 and 22/23

**Figure S15: Effect of age on probability of Influenza A/B test positivity, by influenza vaccination category**

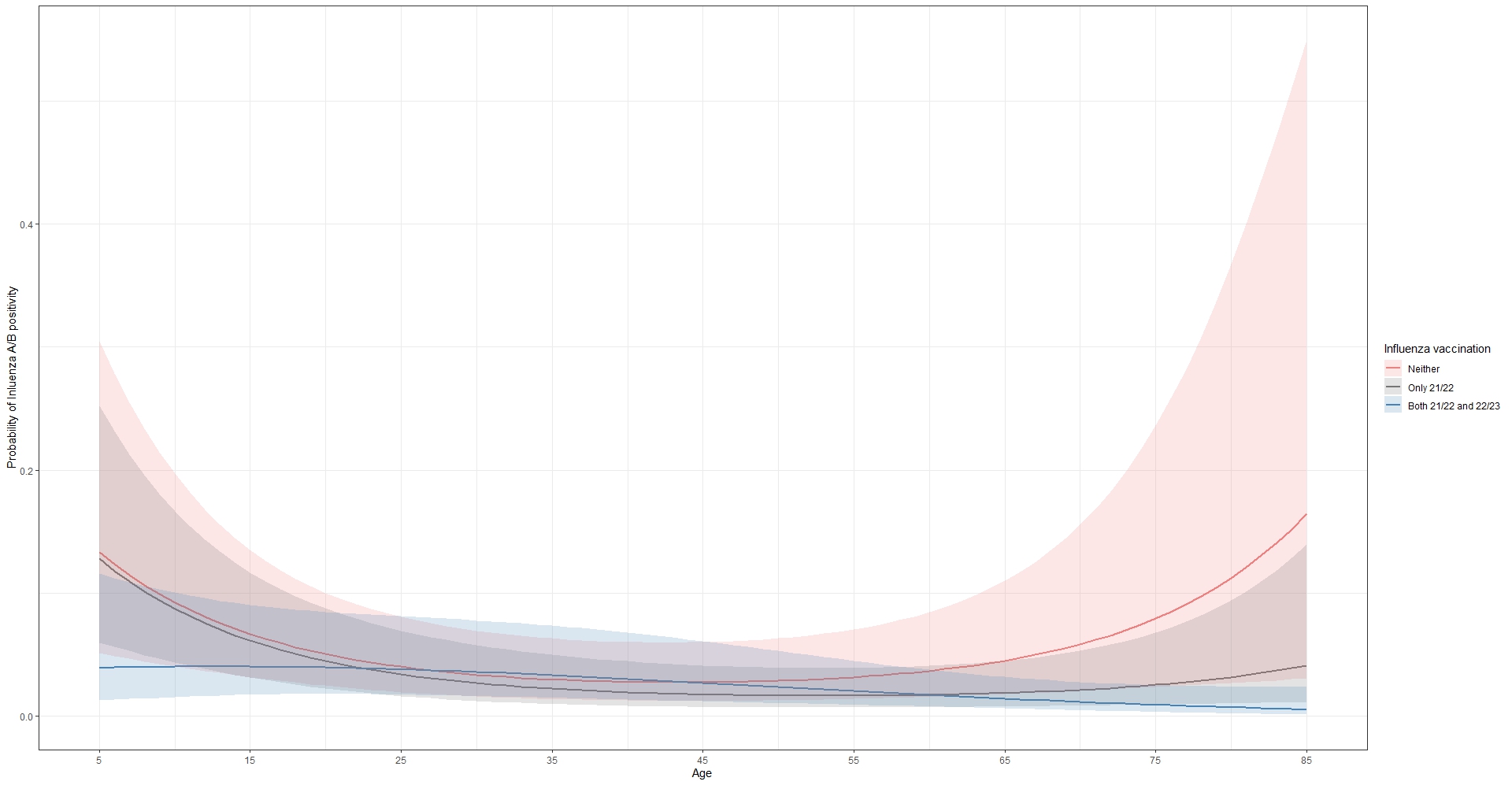

Note: Models included variables seen in Table 2, in addition to smooths for calendar time, days since most recent SARS-CoV-2 vaccination (truncated at 365 days), days since start of most recent SARS-CoV-2 infection episode (truncated at 365 days), and age by flu vaccination status. Probabilities estimated at the following (approximate median for continuous variables): study day 70 (December 18^th^ 2022), Sex=Female, Ethnicity=White, Household size=3 or more, Ever reported long-term health concerns=No, Ever worked in patient-facing health care=No, Prior SARS-CoV-2 infection=Yes, Days since start of most recent SARS-CoV-2 infection = 259, Vaccinated against SARS-CoV-2=Yes, Days since most recent SARS-CoV-2 vaccination=165.

**Figure S16: Effect of Age, calendar time, days since most recent SARS-CoV-2 infection/vaccination on probability of RSV test positivity**
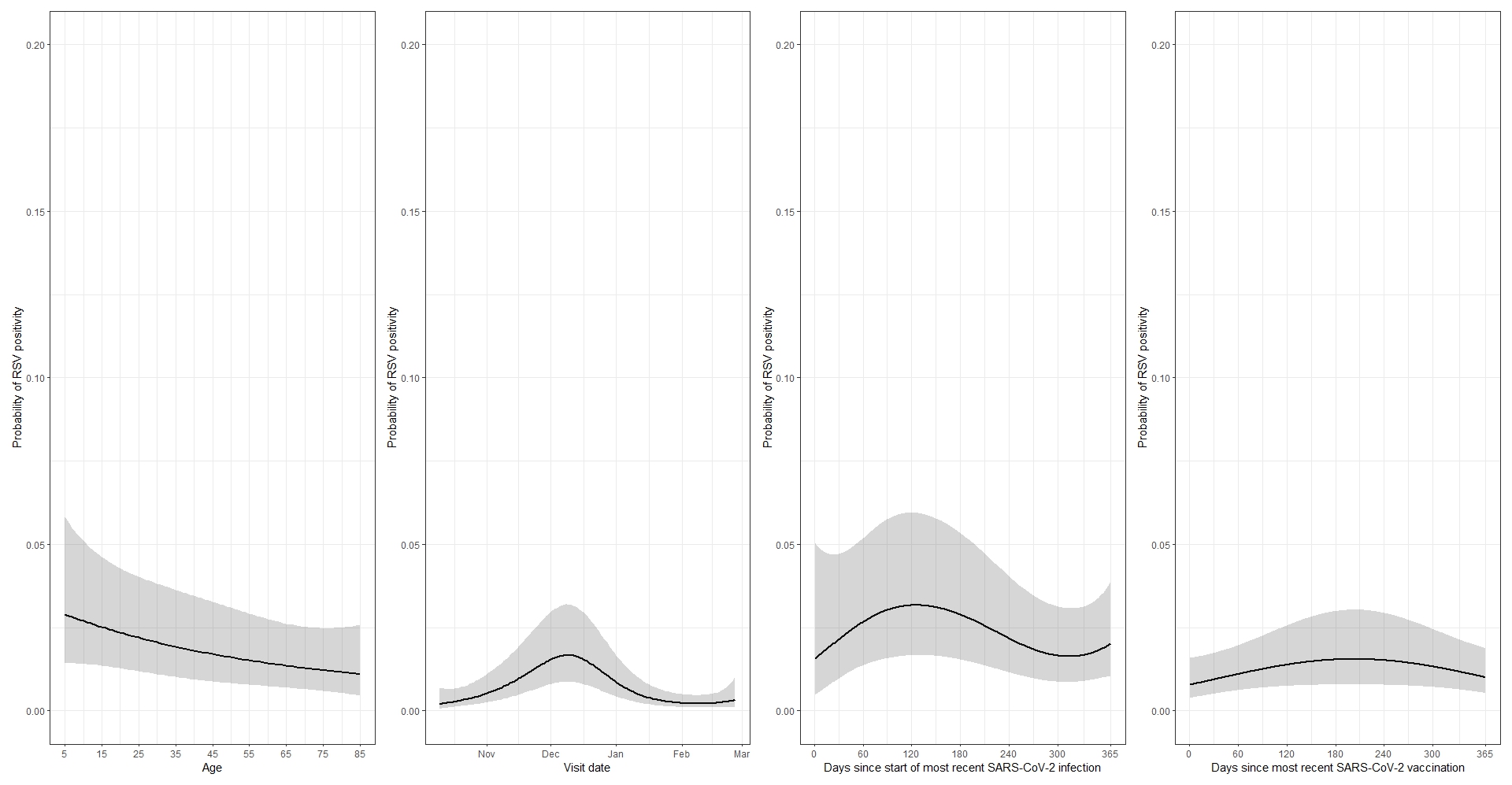

Note: Probabilities estimated at the following (approximate median for continuous variables): study day 70 (December 18^th^ 2022), Sex=Female, Ethnicity=White, Household size=3 or more, Ever reported long-term health concerns=No, Ever worked in patient-facing health care=No, Prior SARS-CoV-2 infection=Yes, Days since start of most recent SARS-CoV-2 infection = 259, Vaccinated against SARS-CoV-2=Yes, Days since most recent SARS-CoV-2 vaccination=165, Flu vaccination=Both 21/22 and 22/23.

**Figure S17: Effect of Age, calendar time, days since most recent SARS-CoV-2 infection/vaccination on probability of SARS-CoV-2 test positivity**

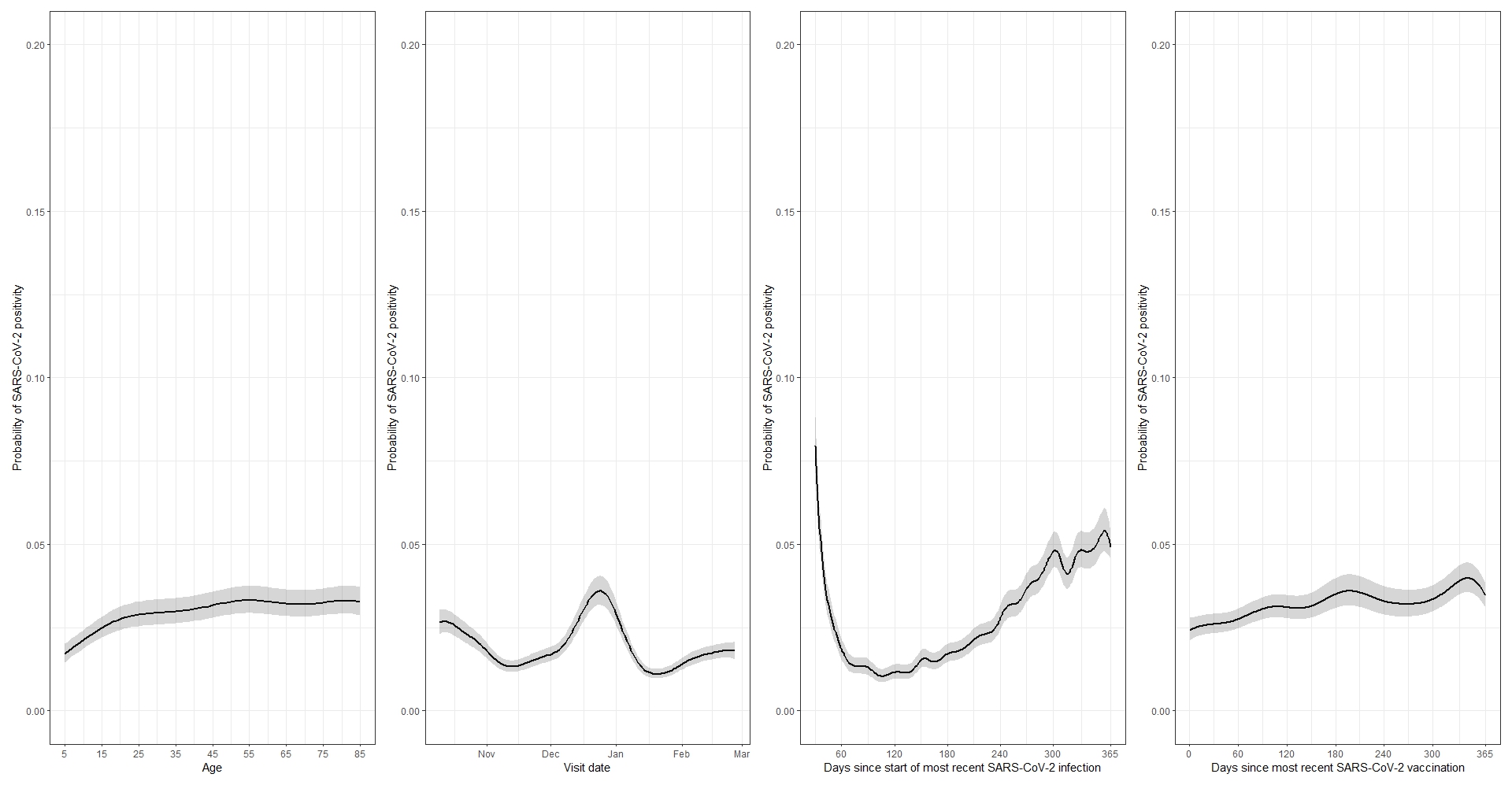

Note: Probabilities estimated at the following (approximate median for continuous variables): study day 70 (December 18^th^ 2022), Sex=Female, Ethnicity=White, Household size=3 or more, Ever reported long-term health concerns=No, Ever worked in patient-facing health care=No, Prior SARS-CoV-2 infection=Yes, Days since start of most recent SARS-CoV-2 infection = 259, Vaccinated against SARS-CoV-2=Yes, Days since most recent SARS-CoV-2 vaccination=165, Flu vaccination=Both 21/22 and 22/23, Upcoming SARS-CoV-2 vaccination in the next 21 days=No. Estimated effects for days since start of most recent SARS-CoV-2 infection are shown after 30 days due to the large estimated probability of a positive test shortly after the start of a recent episode.
